# Genome-resolved metagenomics reveals population-dependent gut microbiome responses to albendazole-ivermectin treatment

**DOI:** 10.64898/2026.11.10.26367728

**Authors:** Javier Gandasegui, Pedro Fleitas, Augusto Messa, Woyneshet Gelaye, Yan Shao, Dawit Degarege, Dereje Bedane, Valdemiro Novela, Áuria de Jesus, Joana Dias, Kirsty Ambridge, Nicole Sevilleno, Anna Bewsher, Terry K. Judah, Inácio Mandomando, Wendemagegn Enbiale, Trevor Lawley, Alejandro Krolewiecki, José Muñoz, Stephen R. Doyle

## Abstract

Soil-transmitted helminth control relies on mass anthelmintic treatment, yet helminth-anthelmintic-microbiome interactions and their role on treatment response remain poorly understood. We characterised the microbiome, resistome, and functional profiles of 858 paired stool metagenomes from 429 children treated with albendazole or a fixed-dose albendazole-ivermectin co-formulation in the ALIVE clinical trial in Ethiopia and Mozambique. Baseline microbiome composition differed more by country than infection status and was not associated with albendazole efficacy or adverse events. Co-formulation treatment, particularly three consecutive daily doses, was associated with depletion of Enterobacteriaceae, predominantly *Escherichia coli*, in Ethiopia, with reductions in *E. coli*-associated resistance genes and metabolic pathways. Genome-resolved analyses of over 22,000 high-quality metagenome-assembled genomes confirmed this depletion across multiple *E. coli* strain-level populations, linking taxonomic, resistome, and functional signals to one ecological event. These findings reveal an off-target effect of combination anthelmintic treatment relevant to large-scale deworming programmes and expand genomic resources for African gut microbiota.

---

Soil-transmitted helminths (STHs) are a group of intestinal nematodes comprising the whipworm *Trichuris trichiura*, the roundworm *Ascaris lumbricoides*, the threadworm *Strongyloides stercoralis*, and the two major human hookworms, *Necator americanus* and *Ancylostoma duodenale^1,2^*. Collectively, they infect nearly one-quarter of the world’s population and cause tremendous morbidity, mainly through chronic malnutrition, malabsorption, anaemia and, in severe cases, intestinal obstruction, resulting in impaired growth and developmental delay^3,4^. To mitigate this burden, World Health Organization (WHO)-promoted control programmes rely primarily on the mass drug administration (MDA) of anthelmintics to at-risk populations in endemic countries, particularly preschool- and school-aged children and women of reproductive age^5^. However, anthelmintics available to treat and control STH infections, mainly albendazole (ALB) and, more recently, in combination with ivermectin (IVM)^6^, show substantial variability in treatment response^7^. Efficacy varies between STH species, being generally lower against *T. trichiura*, intermediate against hookworms and higher against *A. lumbricoides^8^*, but can also differ substantially between and within geographical locations for the same species, particularly for *T. trichiura* and hookworms^9–13^. Among other reasons, the gut microbiome has been proposed to influence treatment response of STH to anthelmintics, as observed for other drugs^14,15^.

As intestinal residents, STH interact closely with the gut microbiome. STH infection has been associated with differences in microbial diversity and composition, although the direction and magnitude of this association vary substantially across helminth species, infection intensity, host age and lifestyle, and geographical setting^16–22^. STH infection has also been linked to shifts in microbial metabolic potential, plausibly reflecting the parasite’s modulation of the intestinal immune and nutritional environment and creating conditions that support mucosal colonisation or persistence within the gut ecosystem^16,20,23,24^.

Considerably less attention has been paid to the direct interplay between anthelmintic treatment, parasite clearance and the gut microbiome. Studies investigating whether the pretreatment microbiome influences drug efficacy have found little evidence of an association with ALB efficacy^25–27^, whereas an exploratory study of ALB-IVM combination treatment reported associations between cohort-specific enterotypes and treatment outcome ^26^. Conversely, longitudinal studies have shown that anthelmintic treatment can shape the gut bacterial composition, although findings have varied across STH species, populations and treatment regimens^27–30^. *In vitro* evidence further suggests that IVM and related compounds can directly inhibit the growth of selected gut bacteria due to antibiotic-like activity^30^. However, these studies have included limited numbers of participants in single cohorts, the reported associations have not been independently validated across geographical settings, and microbiome characterisation has relied predominantly on 16S rRNA gene sequencing or shallow metagenomics, limiting characterisation mainly to taxonomic composition and leaving the mechanisms driving microbiome-worm-treatment interactions largely unknown. In addition, these studies have generally not assessed treatment safety, despite the frequent gastrointestinal disorders associated with anthelmintic drugs and their potential association with the gut microbiome^9,12^.

To address these gaps, we characterised the gut microbiome before and after treatment in the ALIVE clinical trial^12^, which evaluated a new fixed-dose co-formulation (FDC) of ALB and IVM against STH infections, using higher IVM doses than the standard weight-based regimen (200 μg/kg). In this work, we combined read-based analyses with metagenome-assembled genomes (MAGs) to comprehensively characterise the taxonomic composition, resistome, and metabolic and functional profile of the microbiome, defining (i) the baseline characteristics of microbiomes and their association with sociodemographic and infection-related factors, such as STH species and infection intensity, (ii) the interaction between microbiome and treatment efficacy and adverse effects, and (iii) how the different treatment regimens can shape the gut microbiome.

## Results

### Study population and STH infection within the ALIVE clinical trial

The phase 3 of the ALIVE clinical trial evaluated the safety and efficacy of an FDC of ALB and IVM versus ALB monotherapy against *T. trichiura* and hookworms. The FDC was administered either as a single dose (FDCx1) or on three consecutive days (FDCx3). *S. stercoralis*, an initial trial target, was evaluated only as an exploratory outcome due to recruitment difficulties, and *A. lumbricoides* was present only as coinfections. We included participants recruited in Mozambique and Ethiopia with paired pre- and post-treatment stool samples available for microbiome analysis.

After sequencing and quality control (Supplementary Fig. 1A,B), the final dataset comprised 858 samples from 429 participants, including paired pre- and post-treatment samples from 156 participants recruited in Manhiça District, Mozambique, and 273 participants recruited in Bahir Dar, Ethiopia (Fig. 1A). Characteristics of the study population are in Table 1. Among the 156 participants from Mozambique, 107 were infected with *T. trichiura* alone, two with hookworm alone, one had *T. trichiura*-hookworm coinfection, and 46 had *A. lumbricoides-T. trichiura* coinfection (Fig. 1B). In Ethiopia, 230 participants were infected with hookworm alone, 17 with *S. stercoralis* alone, one with *T. trichiura* alone, and 25 had hookworm-*S. stercoralis* coinfection (Fig. 1B). Notably, the participants allocated to FDC arms received higher IVM doses than the standard regimen, with a median IVM dose of 333 μg/kg ranging from 202 μg/kg to 600 μg/kg.

**Fig 1.**
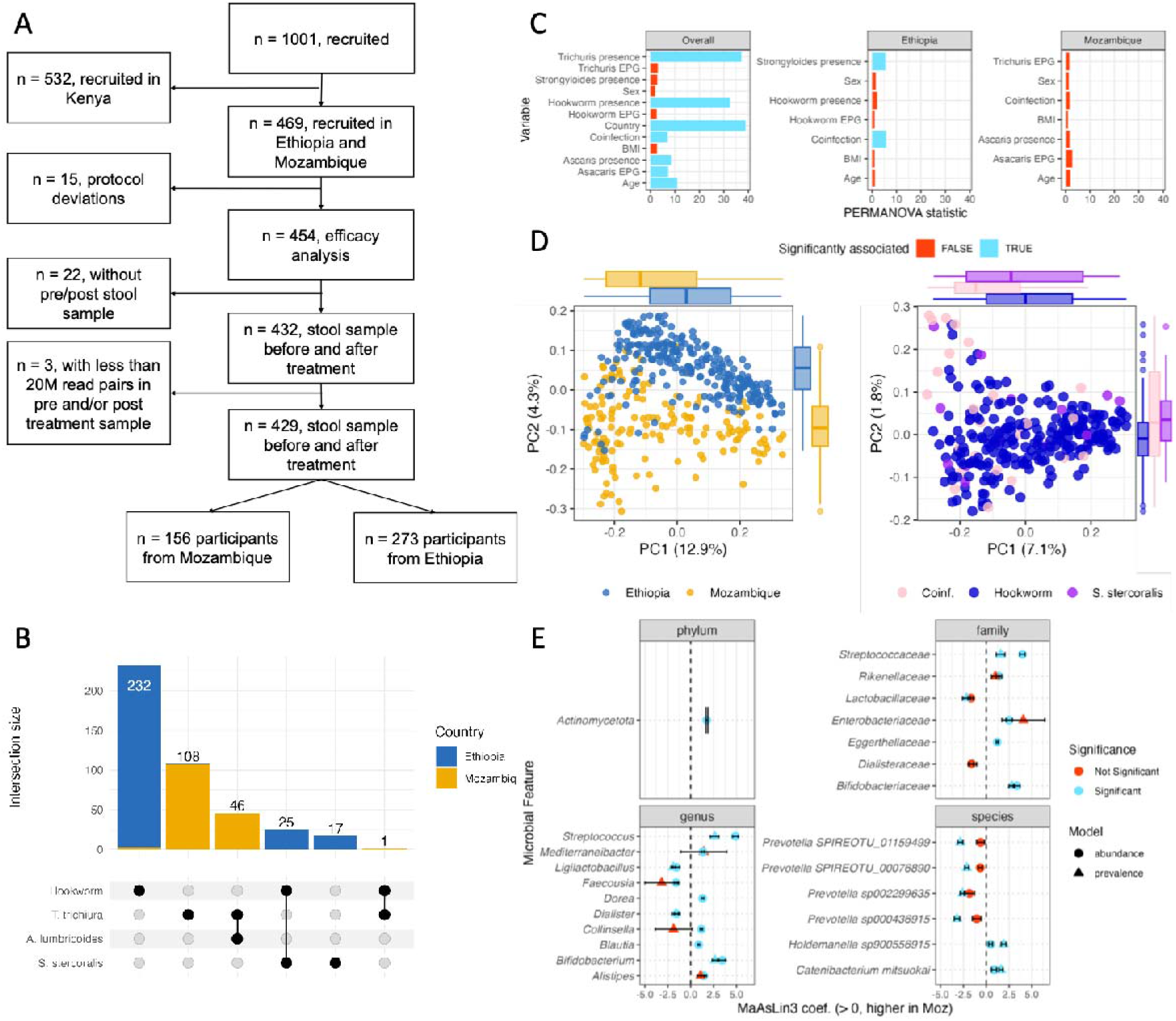
Study population and baseline microbiome characteristics strongly differ between countries. (A) Trial profile showing participant recruitment, exclusion criteria, and the final analytic dataset used for microbiome analyses. (B) UpSet plot showing the intersection sizes of STH infection combinations among the 429 participants, coloured by country. (C) [1] PERMANOVA statistics for the association between baseline gut microbiome composition (Bray–Curtis dissimilarity) and sociodemographic and infection-related variables, shown for the overall cohort and after stratification by country. (D) Principal coordinate analysis (PCoA) of Bray–Curtis dissimilarities in baseline taxonomic composition, coloured by country of recruitment (left) and, within the Ethiopian cohort, by coinfection status (coinfection, hookworm-only, *S. stercoralis*-only; right). (E) Coefficients from MaAsLin3 models testing the association between country of recruitment and microbial feature abundance/prevalence, shown for the 24 bacterial taxa identified as significantly associated with country by both MaAsLin3 and ANCOM-BC2 (consensus features), stratified by taxonomic level (phylum, family, genus, species).

**Table 1.** Characteristics of the study population.

|  | Ethiopia |  |  |  | Mozambique |  |  |  |
|---|---|---|---|---|---|---|---|---|
|  | ALB | FDCx1 | FDCx3 | All | ALB | FDCx1 | FDCx3 | All |
| Participants, n | 86 | 97 | 90 | 273 | 32 | 61 | 63 | 156 |
| Age, mean (SD) | 12.3 (2.85) | 12.1 (3) | 13 (3) | 12.4 (2.9) | 8.8 (3.1) | 9 (2.6) | 8.5 (2.4) | 8.8 (2.6) |
| Female, n (%) | 42 (48.8) | 47 (48.5) | 37 (41.1) | 126 (46.2) | 13 (40.6) | 31 (50.8) | 32 (50.8) | 76 (48.7) |
| BMI, mean (SD) | 16.4 (2.1) | 16.4 (2.6) | 16.6 (2.3) | 16.5 (2.3) | 15.7 (3.1) | 15.3 (2.3) | 15.2 (2.1) | 15.3 (2.4) |
| Tt infected, n | 0 | 1 | 0 | 1 | 31 | 60 | 63 | 154 |
| Tt EPG, mean (SD) | 0 (0) | 1 (6) | 0 (0) | 0.4 (3.6) | 631 (1359) | 517 (1516) | 735 (1465) | 628 (1459) |
| Hwk infected, n | 83 | 88 | 84 | 255 | 1 | 2 | 0 | 3 |
| Hwk EPG, mean (SD) | 291 (1072) | 288 (966) | 372 (917) | 316 (982) | 6 (36) | 3 (23) | 0 (0) | 2.4 (21.7) |
| Al infected, n | 0 | 0 | 0 | 0 | 13 | 16 | 17 | 46 |
| Al EPG, mean (SD) | 0 (0) | 0 (0) | 0 (0) | 0 (0) | 4522 (8037) | 2578 (6150) | 3171 (6898) | 3216 (6864) |
| Ss infected, n | 10 | 16 | 16 | 42 | 0 | 0 | 0 | 0 |

BMI, body mass index; Tt, *T. trichiura*; Hwk, hookworm; Al, *A. lumbricoides*; Ss, *S. stercoralis*; EPG, eggs per gram of stool; SD, standard deviation.

### Baseline microbiome characterisation reveals differences between geographic locations and coinfected participants

Overall taxonomic microbiome structure was most strongly associated with the country, although age and all infection-related variables were also significant in the full cohort (Fig. 1C,D). However, STH species were highly unevenly distributed between study sites, and participant age also differed between countries (Fig. 1B and Table 1); we therefore stratified subsequent analyses by country. After stratification, only *S. stercoralis* infection in Ethiopia, particularly among participants coinfected with hookworm, remained significantly associated with taxonomic variation (Fig. 1C,D; Supplementary Tables S1). Shannon-index alpha diversity did not differ significantly according to any of the baseline variables evaluated (Supplementary Tables S1).

We next examined which bacterial taxa differed between countries, and between mono- and coinfected participants only in Ethiopia. Using MaAsLin3^31^ and ANCOM-BC2^32^, 24 taxonomic features differed significantly between countries by both differential-abundance (DA) testing methods (Fig. 1E, and Extended Data Figs. 1 and 2). By contrast, no taxonomic features were associated with coinfection status in Ethiopia using both methods, although MaAsLin3 alone identified five (Extended Data Fig. 1), including depletion of *Bacteroidota*, *Bacteroidaceae,* and *Prevotella* in coinfected individuals.

**Fig 2.**
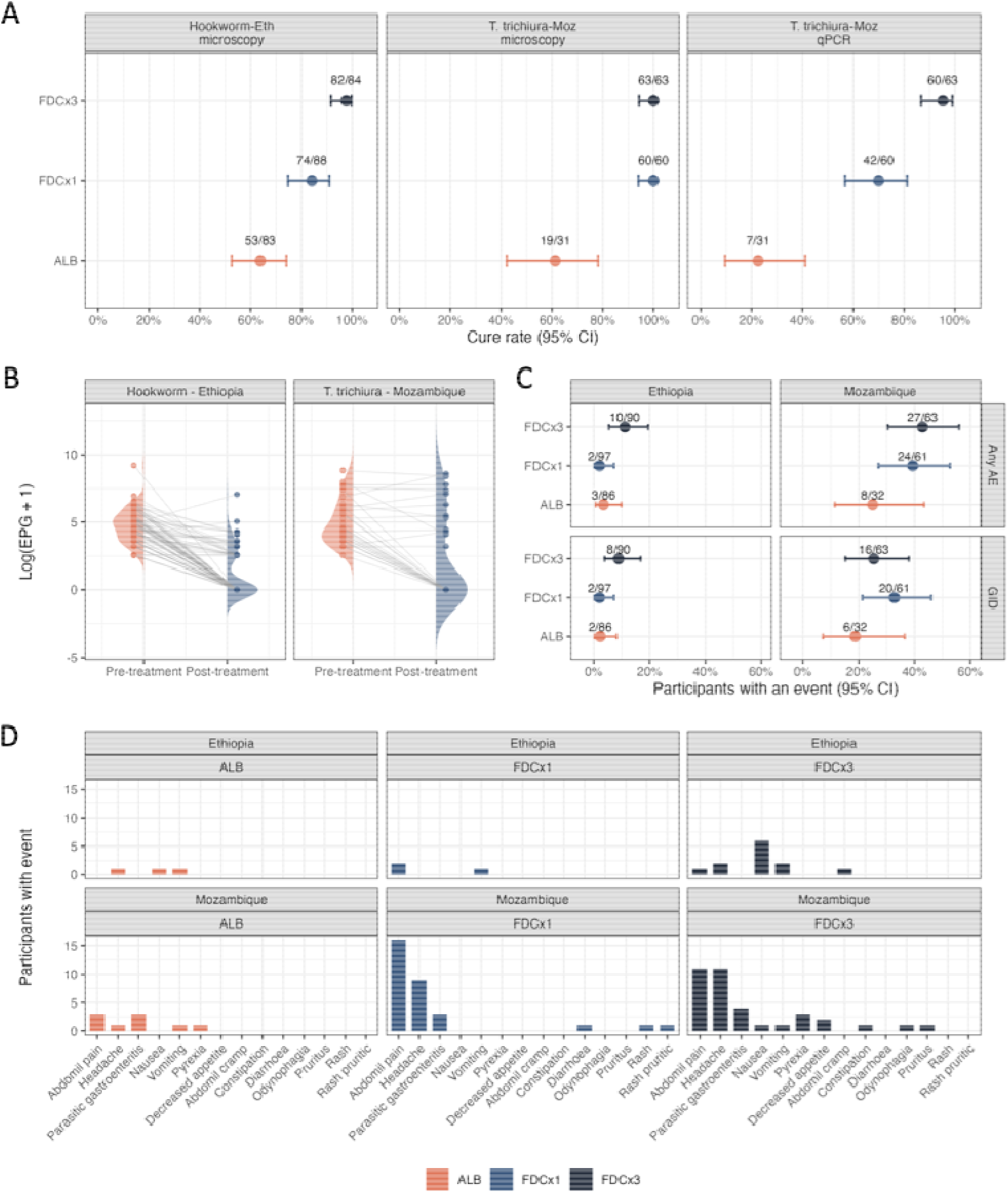
Treatment efficacy and safety across regimens and countries. (A) Cure rates (95% CI) for hookworm in Ethiopia (microscopy) and T. trichiura in Mozambique (microscopy and qPCR), by treatment arm. Point labels indicate the number of participants cured relative to the number infected at baseline. (B) Paired individual egg counts (log(EPG + 1)) before and after treatment among ALB-arm participants infected with hookworm (Ethiopia) or T. trichiura (Mozambique), with half-violin density distributions shown for the pre- and post-treatment timepoints and grey lines connecting each participant’s paired values. (C) Proportion of participants reporting any adverse event (AE) (top) or a gastrointestinal disorder (GID; bottom) by country and treatment arm. (D) Number of participants reporting each specific adverse event term, stratified by country and treatment arm.

Baseline antimicrobial resistance genes (ARGs) and functional read-based compositional profiles showed patterns similar differential patterns to taxonomy (Extended Data Fig. 3A-D): in the full cohort, age and all evaluated infection-related variables were significantly associated with both profiles, with the country of origin again showing the strongest association. After stratification, hookworm-*S. stercoralis* coinfection in Ethiopia and *T. trichiura*-*A. lumbricoides* coinfection in Mozambique remained significantly associated with resistome and functional composition (Extended Data Fig. 3A,B). Resistome alpha diversity differed significantly between countries (Extended Data Fig. 3E), being higher in Mozambique for both set of data, and, after stratification, between mono- and coinfected participants in Ethiopia only (Extended Data Fig. 3F).

DA analyses identified 33 ARGs and 170 metabolic pathways that differed significantly between countries, including depletion of the *Bacteroidales*-associated ARGs in Ethiopia and marked enrichment of aerobic-respiration pathways in Mozambique (Extended Data Fig. 4). In contrast, features associated with coinfection status within countries were identified only by MaAsLin3 (Extended Data Figs. 1). These findings demonstrate pronounced differences in the taxonomic, resistome and functional profiles of the gut microbiome between Mozambican and Ethiopian participants, with substantially weaker within-country coinfection associations. Subsequent analyses were therefore stratified by country. Complete statistical results are in Supplementary Tables S1-S3 (diversity) and S4-S6 (differential abundance).

### Pretreatment gut microbiomes are not associated with drug efficacy or safety

We next assessed whether pretreatment microbiome composition was associated with treatment efficacy, initially defined as cure using microcopy. As previously reported^12^, FDCx1 and FDCx3 achieved high efficacy, particularly against *T. trichiura*; in our microbiome cohort, both regimens achieved cure rates of 100% in Mozambique and at least 80% in Ethiopia (Fig. 2A). Few participants remained infected after treatment, precluding adequately powered cured-vs-non-cured comparisons in those arms, so we restricted the primary efficacy analyses to participants receiving ALB alone — those infected with *T. trichiura* in Mozambique and hookworm in Ethiopia.

Baseline alpha/beta diversity and differential abundance (by either MaAsLin3 or ANCOM-BC2) showed no significant association with treatment outcome (Supplementary Table S7). Secondary efficacy endpoints, individual egg reduction rate (ERRi)^33^ (Fig. 2B) and qPCR-defined cure only for *T. trichiura^34^* (Fig 2A), likewise showed no significant microbiome associations (Supplementary Table S7). Because IVM may provide limited additional efficacy over ALB against hookworm^10^, implying FDCx1 and ALB could be equivalent, we also compared baseline microbiome composition between cured and non-cured participants in the merged ALB-FDCx1 arms in Ethiopia; this again identified no significant associations (Supplementary Table S7). Overall, these analyses provided no evidence that baseline taxonomic composition, resistome or functional potential was associated with ALB efficacy.

We also assessed whether the pretreatment microbiome was associated with treatment safety, as measured by adverse event (AE) occurrence. AEs were most frequent in the FDCx3 arm, followed by FDCx1 and ALB (Fig. 2C,D), and occurred most often among Mozambican participants (Fig. 2C); gastrointestinal disorders (GIDs) — particularly abdominal pain and headache — were the most frequently reported (Fig. 2D). Within each treatment arm, we first compared baseline taxonomic composition and functional profile of participants with and without any AE, and separately for treatment-related GIDs given their potential link to the intestinal microbiome; neither analysis identified significant associations in any arm (Supplementary Table S7). Pooling FDCx1 and FDCx3 participants to increase power likewise identified no significant associations for overall AEs or GIDs (Supplementary Table S7). Collectively, these analyses provided no evidence that pretreatment microbiome characteristics were associated with AE occurrence.

### ALB-IVM FDC treatment shapes the gut microbiome composition, resistome and functional profile

We assessed the impact of treatment on gut microbiome taxonomic composition, resistome, and functional profile, comparing samples before and after treatment, stratified by country and treatment arm. At the taxonomic level in Mozambique, we observed no significant differences in alpha or beta diversity after treatment (Supplementary Table S8) and, although MaAsLin3 identified several differentially abundant taxa and metabolic pathways, ANCOM-BC2 confirmed none (Extended Data Fig. 5).

By contrast, FDC treatment in Ethiopia was associated with a decrease in *Enterobacteriaceae* in both FDCx1 and FDCx3, and *Escherichia* and *E. coli* in FDCx3 (Fig. 3A). Reductions were also observed in Mozambique but were not significant by either method (Fig. 3A). No correlation was observed between individual ALB or IVM doses and these reductions (Supplementary Fig. 2). Repeating the analysis among only ALB-arm participants who achieved cure — to test whether parasite clearance itself, rather than non-cured participants masking an effect, drove the *Enterobacteriaceae* decrease — identified significant taxa only by MaAsLin3, with *Enterobacteriaceae* and its members not significantly reduced (Supplementary Table S9). In the Ethiopian FDCx3 arm, the alpha diversity of both resistance genes and metabolic pathways (Shannon index) decreased significantly after treatment (Extended Data Fig. 6A). Overall composition of the resistome and pathway profiles (PERMANOVA) also shifted after both FDCx1 and FDCx3 (Extended Data Fig. 6B; Supplementary Table S8), whereas taxonomic diversity and composition did not show significant changes change.

**Figure 3.**
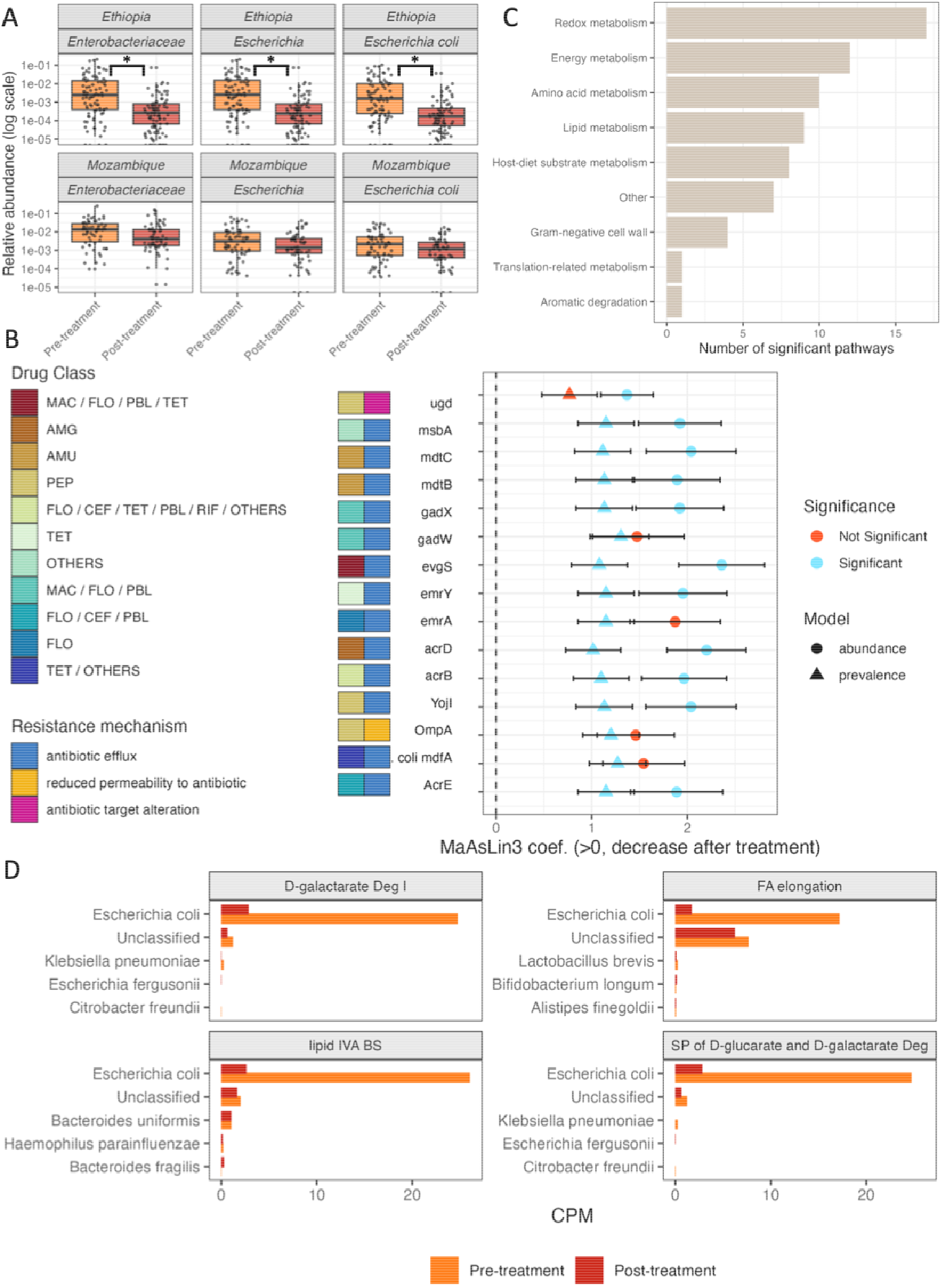
FDCx3 treatment is associated with changes in gut microbiome taxonomic, resistome and functional profiles. (A) Relative abundance of *Enterobacteriaceae, Escherichia* and *Escherichia coli* before treatment and after treatment among participants receiving FDCx3 treatment in Ethiopia and Mozambique; *: significant association by MaAsLin3 and ANCOM-BC2. (B), MaAsLin3 coefficients for the 15 ARGs identified as differentially abundant by both MaAsLin3 and ANCOM-BC2 following FDCx3 treatment in Ethiopia. Positive coefficients indicate a reduction after treatment. Error bars indicate ±1 standard error. Tiles indicate the associated antibiotic classes and resistance mechanisms. (C), Functional classification of the 69 metabolic pathways that decreased after FDCx3 treatment in Ethiopia and were identified as significant by both MaAsLin3 and ANCOM-BC2. Bars show the number of pathways assigned to each functional category. (D), Mean species-stratified HUMAnN abundance, expressed as copies per million (CPM), before and after FDCx3 treatment for the five principal taxonomic contributors—including unclassified contributions—to four of the most abundant pathways. (B): AMG, aminoglycoside; AMU, aminocoumarin; CEF, cephalosporin; FLO, fluoroquinolone; MAC, macrolide; PBL, penicillin β-lactam; PEP, peptide antibiotic; RIF, rifamycin; TET, tetracycline. (D): Deg, degradation; FA, fatty acid; BS, biosynthesis; SP; super-pathway.

DA analyses likewise identified features consistently detected by both methods in the FDCx1 arm and, more prominently, the FDCx3 arm (Extended Data Fig. 5). Complete results are in Supplementary Tables S9-S11 (taxa, ARGs, and pathways, respectively). For the resistome, both methods identified 15 ARGs conferring resistance to various antibiotic families as significantly less abundant after FDCx3 (Fig. 3B, Extended Data Fig. 5 and Supplementary Table S10,11). Pathway DA showed more changes after FDCx3 than after FDCx1: 69 pathways were significantly lower after FDCx3 (by both methods) versus 2 after FDCx1 treatment (Fig. 3C and Extended Data Fig. 5). Species-stratified HUMAnN profiles showed that *E. coli* was the predominant contributor to most of these pathways (Extended Data Fig. 7), with pathway-specific contributions substantially lower after treatment (Fig. 3D, four most abundant pathways), indicating this reduction was largely driven by *E. coli* depletion.

### Genome-resolved analyses confirm broad, strain-nonspecific depletion of E. coli and its carried ARGs

To further characterise these changes in the microbiome, we reconstructed MAGs from all pre- and post-treatment samples. Across the 858 stool metagenomes, we recovered 60,738 MAGs and retained 22,032 high-quality MAGs for downstream analyses after quality filtering. Dereplication at 95% average nucleotide identity (ANI) grouped these MAGs into 1,372 representative species-level genomes, comprising 1,366 bacterial (Extended Data Fig. 8A) and six archaeal genomes (Extended Data Fig. 8B). Of the bacterial representatives, 516 were recovered in both countries, while 528 and 324 were detected exclusively in Ethiopia and Mozambique, respectively (Extended Data Fig. 8C–E). Among the 22,032 high-quality MAGs, 518 could not be assigned to a species by GTDB-Tk and were grouped into 161 representative genomes. Of these, 158 could be taxonomically assigned at the genus level, and three only at the family level (Extended Data Fig. 9), likely representing poorly characterised or novel species-level populations.

We next investigated whether the treatment-associated reduction in *Enterobacteriaceae*, particularly *E. coli*, reflected the depletion of specific phylogenetic lineages or strain-level populations. Comparing read-based abundance with the recovery and distribution of *Enterobacteriaceae* MAGs (Fig. 4A) showed that composition differed markedly between countries: *E. coli* dominated in Ethiopia, whereas Mozambique contained a prominent, treatment-stable *Klebsiella pneumoniae* population alongside an additional *E. coli* population and minority taxa uncommon in Ethiopia (Fig. 4A). Dereplicating *Enterobacteriaceae* MAGs at 99% ANI yielded 50 strain-level clusters (Supplementary Fig. 3A), 31 of which were *E. coli* (Fig. 4B). Two *E. coli* clusters were substantially more prevalent, with no evidence of country-specific populations (Supplementary Fig. 3B); major strains showed similar treatment responses within each country, with the clearest reductions following FDC in Ethiopia (Fig. 4C), indicating the depletion was not driven by loss of one dominant strain but reflected a broader reduction in *E. coli*.

**Figure 4.**
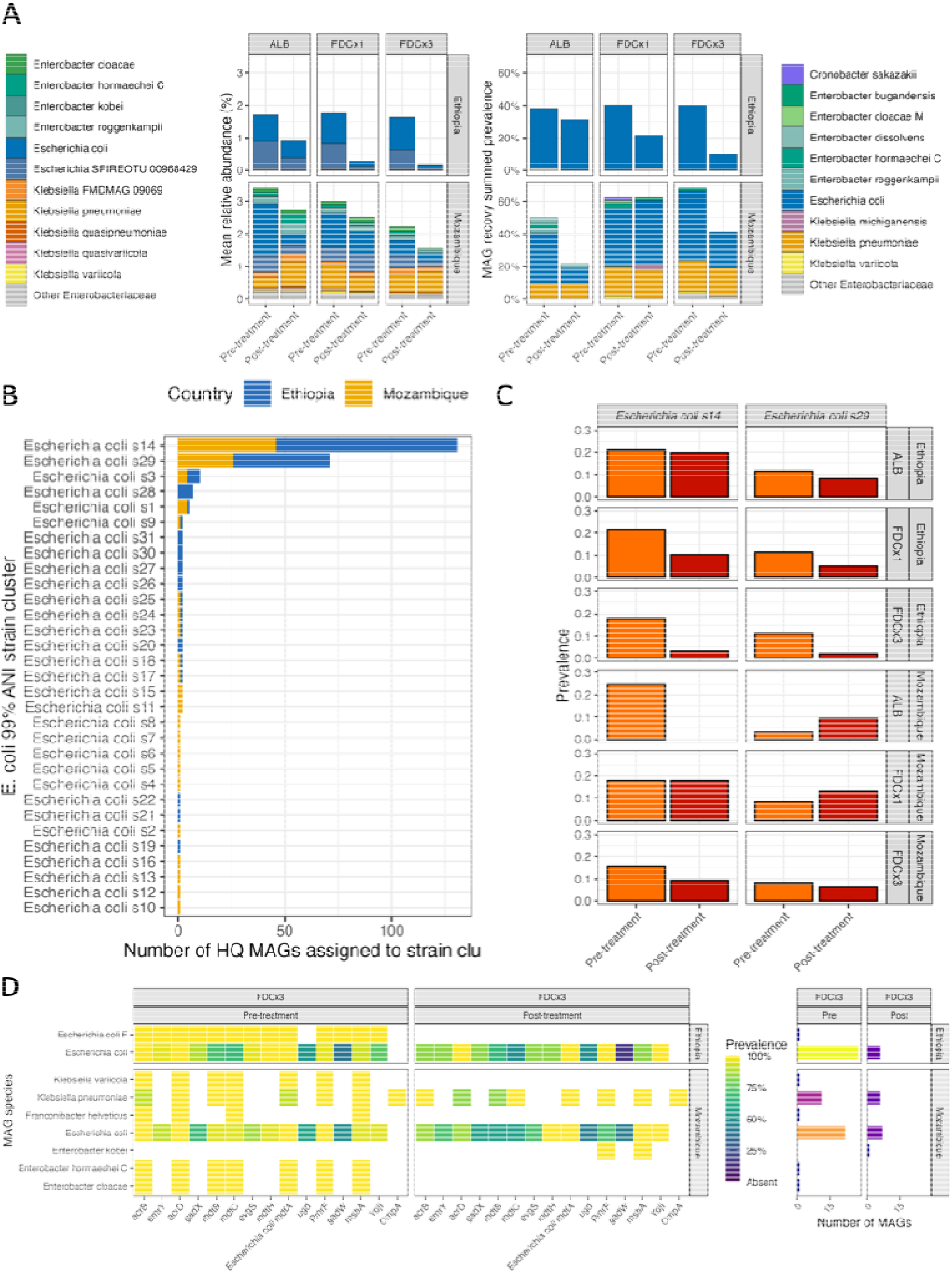
Genome-resolved analyses support a broad depletion of *Escherichia coli* and associated antimicrobial resistance genes following FDC treatment. (A) Species composition of *Enterobacteriaceae* before and after treatment, stratified by country and treatment arm. Stacked bars show read-based mean relative abundance estimated using Sylph (left) and species-specific MAG recovery prevalence (right). (B) Number of high-quality *E. coli* MAGs assigned to each of the 31 strain-level clusters obtained by dereplication at 99% average nucleotide identity (ANI), coloured by country. (C) Recovery prevalence of the two dominant *E. coli* strain-level clusters, s14 and s29, before and after treatment, stratified by country and treatment arm. (D) Distribution of the 15 antimicrobial resistance genes (ARGs) that decreased significantly after FDCx3 treatment in Ethiopia among high-confidence *Enterobacteriaceae* MAGs recovered before and after FDCx3 treatment. Heatmap colours indicate the proportion of MAGs assigned to each species that carried the corresponding ARG; white denotes absence of the MAG. Adjacent bars show the number of MAGs contributing to each species-level estimate. HQ, high quality; MAG, metagenome-assembled genome.

We also characterised the resistome of reconstructed MAGs using RGI/CARD, enabling direct comparison with the read-based resistome analyses. Overall, 14,719 high-quality MAGs harboured at least one ARG (mean 3.85 ARGs/MAG). The 15 ARGs significantly reduced after FDCx3 in the read-based analysis were carried exclusively by *Enterobacteriaceae* MAGs, almost all specifically by *E. coli* (except *OmpA*), with no significant variation after FDCx3 treatment (Fig. 4D). Overall, FDC treatment, particularly three consecutive doses, was associated with a broad, strain-nonspecific depletion of *E. coli* in Ethiopia, accompanied by depletion of *E. coli*-carried ARGs and metabolic pathways.

### Metabolic pathway changes converge across countries and regimens despite limited taxonomic and resistome overlap

Because requiring concordant MaAsLin3-ANCOM-BC2 significance might be conservative, we also examined features significant by MaAsLin3 alone, intersected between countries within each arm, and between FDCx1 and FDCx3 within each country. No features were shared between countries in the ALB arm, and taxonomic/resistome responses to the FDC regimens also showed limited overlap between countries (Fig. 5A,B). *Catenibacterium* decrease was the only effect associated with FDCx1 treatment in both Ethiopia and Mozambique, though with small effect estimates and no ANCOM-BC2 support (Supplementary Table S8). No taxa or ARGs were shared between FDCx1 and FDCx3 in Mozambique (Fig. 5A,B). Within Ethiopia, *Enterobacteriaceae*, *Escherichia* and *E. coli* were the only taxonomic features associated with both FDC regimens (Fig. 5A), reinforcing the treatment-associated reduction in this bacterial lineage in this country. For the resistome, all six ARGs associated with FDCx1 were also associated with FDCx3; four — *tet(W), tet(40), tet(O/32/O)* and *tet(O/M/O)* — showed concordant increases after both regimens (Fig. 5C, Supplementary Table S10), distributed across a broad range of bacterial taxonomic groups when mapped to high-quality MAGs (Fig. 5D).

**Figure 5.**
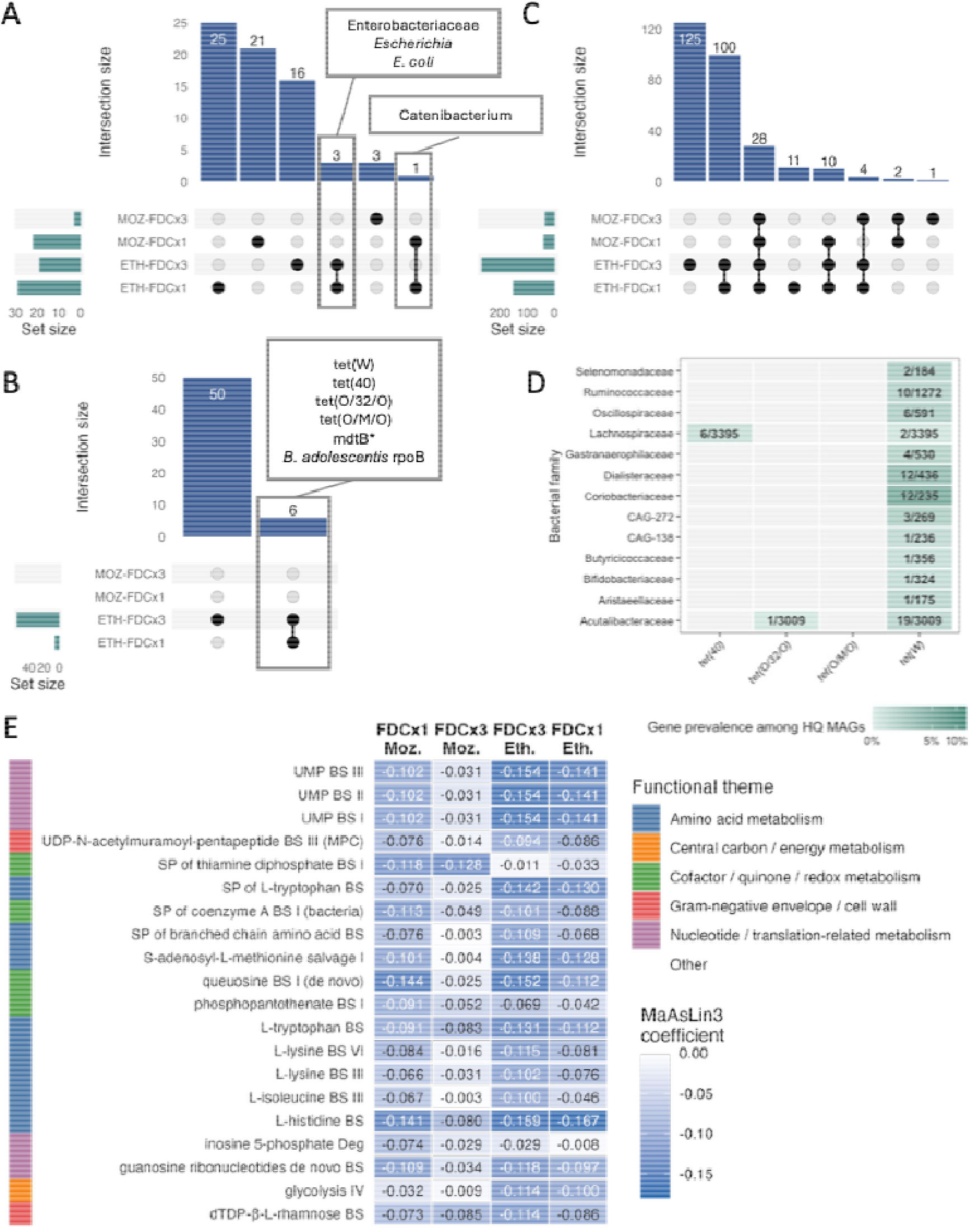
Shared taxonomic, resistome and functional responses to fixed-dose albendazole–ivermectin treatment across regimens and countries. (A–C), UpSet plots showing the intersections of taxonomic features (A), antimicrobial resistance genes (ARGs; B) and metabolic pathways (C) identified as significantly different between pretreatment and post-treatment samples by MaAsLin3 in Ethiopia and Mozambique. (D), Prevalence of tet(W), tet(40), tet(O/32/O) and tet(O/M/O) among high-quality metagenome-assembled genomes (MAGs), stratified by bacterial family. Colour intensity represents the proportion of high-quality MAGs within each family carrying the indicated gene, and values show the number of gene-positive MAGs divided by the total number of high-quality MAGs assigned to that family. Only families carrying at least one of the four genes are shown. (E), MaAsLin3 coefficients for the 20 metabolic pathways that increased concordantly following both FDC regimens in Ethiopia and Mozambique. Rows represent pathways and columns represent country-by-treatment comparisons; negative coefficients indicate greater pathway abundance after treatment because the model contrast was pretreatment versus post-treatment. The coloured row annotation indicates the assigned functional category. HQ, high quality; * in panel (B), means that the gene mdtB was also significant by ANCOM-BC2 in the FDCx3 arm.

In contrast, metabolic pathways showed substantially greater cross-country overlap: 28 pathways were detected in all four country-by-treatment comparisons, of which 20 increased concordantly across both countries and FDC regimens (Fig. 5E). These comprised pathways spanning amino acid metabolism (n=8), nucleotide/translation-related metabolism (n=5), cofactor/quinone/redox metabolism (n=4), cell-wall metabolism (n=2) and central carbon metabolism (n=1). Species-stratified HUMAnN profiles indicated this increase was distributed across multiple bacterial contributors rather than one species (Extended Data Fig. 10), consistent with functional redundancy such that a community-wide shift in growth state or niche occupancy can register functionally without a corresponding taxon-level signal. Together, these findings suggest that, beyond the pronounced depletion of *E. coli* in Ethiopia, FDC treatment may induce a weaker but more broadly distributed ecological response, reflected in relative increases in tetracycline-resistance genes across diverse taxa in Ethiopia and in metabolic pathways shared across regimens in both countries.

## Discussion

In this study, we provide a comprehensive assessment of interactions among STH infection, anthelmintic treatment, and the gut microbiome in a large multicentre clinical trial. Three major findings emerged. First, geographical origin was the strongest determinant of microbiome variation, substantially outweighing associations with infection status or other baseline variables. Second, we found no evidence that baseline microbiome composition, resistome, or functional profile was related to albendazole efficacy or to the occurrence of adverse events. Third, combination treatment with ALB and high doses of IVM was associated with changes in the gut microbiome, particularly after three consecutive doses, that in Ethiopia was characterised by depletion of *Enterobacteriaceae* and *E. coli* and accompanied by reductions in *E. coli*-associated metabolic pathways and ARGs.

The influence of helminth coinfection and microbiome characteristics, as we observed in our baseline analysis, has been described previously^16–18,22^, although the specific features suggest more than a generic consequence of harbouring multiple helminth species. In Ethiopia, we found a weak coinfection signal plausibly attributable to *S. stercoralis*, which has been specifically linked to mucin-degrader enrichment and altered short-chain fatty acid metabolism during chronic infection ^35,36^. Similar country-of-origin effects have been reported in other microbiome clinical studies, highlighting one of the major challenges of identifying microbiome biomarkers in geographically diverse populations^37,38^. This challenge is further exacerbated in African populations, which remain substantially underrepresented in public microbiome databases and large-scale reference studies^39–41^. By characterising two independent African cohorts, our study both demonstrates the importance of geographical context in microbiome associations and contributes to thousands of African MAGs, expanding genomic reference resources for this underrepresented population.

We found no evidence that baseline microbiome composition was associated with ALB treatment efficacy, which is relevant for drug pharmacokinetics since gut bacteria can metabolise this drug into its active compounds^42^, potentially influencing efficacy. No differential-abundance test identified a taxon, ARG, or pathway significantly different between cured and non-cured participants, consistent with previous studies ^25,26^. Nonetheless, ALB may not be an entirely inert comparator: independent studies have reported ALB-associated microbiome changes even in uninfected individuals ^28,43^, warranting some caution in treating it as a fully neutral comparator for the FDC-associated signal described below.

FDC treatment produced clear microbiome changes in Ethiopia, more pronounced after three consecutive doses. A recent study reported dose-associated microbiome alterations stratified post hoc by absolute IVM milligram dose^29^; however, because IVM was dosed to a constant weight-adjusted target (200 μg/kg), absolute dose should chiefly reflect body weight rather than IVM intestinal concentration derived from per-kilogram dosage. Our fixed-dose FDC regimen, by contrast, produced genuine inter-individual variation in weight-adjusted exposure (202–600 μg/kg), offering a more direct test of a true dose-response relationship: we found no correlation between individual weight-based estimated IVM or ALB doses and the depletion of Enterobacteriaceae or *E. coli*. However, we observed greater changes in taxonomic composition, resistome, and functional potential after repeated administration, suggesting that the stronger microbiome effect may be associated with cumulative exposure rather than differences in instantaneous intestinal IVM concentration. The clearest FDC-associated signal was a broad depletion of *Enterobacteriaceae*, predominantly *E. coli*, in Ethiopia. Because this family includes both commensal organisms and important opportunistic pathogens and reservoirs of antimicrobial resistance^44^, the clinical significance of its depletion remains uncertain. The parallel loss of ARGs and their bacterial carriers suggests resistome changes primarily reflected *E. coli* depletion rather than selection for resistance^45^. Indeed, most of the 15 ARGs identified are intrinsic, chromosomally-encoded efflux-pump components present in essentially all E. coli genomes rather than acquired resistance determinants — consistent with an independent ALB-IVM trial reporting that ARG dynamics largely mirrored host MAG abundance, with limited gene-specific selection^29^. By contrast, our taxon-independent increase in tetracycline-resistance genes in both FDC arms (*tet(W), tet(40), tet(O/32/O), tet(O/M/O)*) runs counter to a reported decrease in related genes after single-dose ALB-IVM^29^, possibly reflecting cumulative-versus single-dose exposure.

Although direct antibacterial activity of IVM has been demonstrated *in vitro^30^*, our findings do not establish such an effect *in vivo,* and recent evidence suggests this may be specifically unlikely for *E. coli*: in a panel of gut isolates repeatedly exposed to IVM and moxidectin, *E. coli* was the only species showing no clear phenotypic or genomic shift, attributed to limited drug access across its Gram-negative envelope and consistent with reported *E. coli* insensitivity to macrolide antibiotics ^30^. This aligns with our own strain-level data, where depletion was broadly distributed across 31 *E. coli* clusters with no evidence of a selective sweep (Fig. 4B,C), more consistent with indirect, ecological depletion than direct selection. Notably, the same study found downregulated multidrug-efflux gene expression (*mdtI*, *mdtJ*, *ydhC*) upon exposure, raising the possibility that some ARG depletion reflects altered expression rather than abundance alone. Alternative mechanisms are also plausible: helminths modulate both the intestinal microbiota and host immune responses^46,47^, and successful treatment could progressively reshape the gut environment as mucosal inflammation resolves^28^, although the absence of significant features in only-cured individuals in the ALB arm in Ethiopia suggests that this cannot be the only mechanism.

Hookworm-induced blood loss may also increase luminal iron and heme, substrates *E. coli* exploits efficiently via siderophore and heme-uptake systems^48^, plausibly explaining its predominance in hookworm-dominated Ethiopia relative to the stable *K. pneumoniae* population in *T. trichiura*-dominated Mozambique, and the corresponding absence of a treatment effect there. Finally, IVM has documented host-directed anti-inflammatory properties^49^ that, together with parasite clearance, could explain the pronounced effect after repeated FDCx3 administration without requiring direct antibacterial activity.

The ALIVE trial was primarily designed to evaluate the safety and efficacy of the FDC compared with ALB^12^, which imposed several constraints on the microbiome analyses. Competitive recruitment facilitated participant enrolment^50^ but resulted in each country being dominated by its locally prevalent STH species, limiting our ability to fully distinguish geographical from parasite-specific effects. Although sampling 21 days after treatment was optimal for assessing anthelmintic efficacy^51^, additional time points, such as 24- or 48-hour stool collection, would have helped determine whether the change was immediate, suggesting or excluding cytotoxicity as observed for antibiotics^52–54^. Similarly, measuring acute inflammation or haematological markers would have helped clarify the mechanisms underlying these microbiome alterations^55,56^. Despite these limitations, deep shotgun metagenomics substantially strengthened our analysis compared with previous shallower or 16S-based approaches, which typically offer only genus-level resolution without detailed access to functional or resistome composition^57^. For example, genome-resolved analyses let us trace ARG changes to their bacterial hosts and confirm *E. coli* depletion at the strain level, demonstrating the value of this approach for interpreting functional and resistome shifts in clinical trials.

In conclusion, our study shows that ALB–IVM FDC treatment is associated with changes in the gut microbiome beyond the clearance of target parasites, particularly after repeated administration, including broad depletion of *E. coli* and a resistome shift marked by both loss of *E. coli*-linked resistance genes and broader enrichment of tetracycline-resistance determinants across unrelated taxa. As ALB–IVM combination therapy moves towards extensive use in STH control programmes, understanding its off-target effects on the intestinal microbiome, and distinguishing transient ecological perturbations from clinically meaningful consequences, will become increasingly important.

## Methods

### Study design and participants

Samples were collected within the ALIVE trial (ClinicalTrials.gov NCT05124691)^12^, an adaptive phase 2/3, single-blind (outcome-assessor-blinded), randomised, multicentre, parallel-group, active-controlled superiority trial evaluating a fixed-dose co-formulation (FDC) of ALB (400 mg) and ivermectin (9 or 18 mg) against ALB monotherapy (400 mg) for the treatment of *T. trichiura*, hookworm, and *S. stercoralis* infections. The trial was conducted across 15 schools in Kwale County (Kenya), Bahir Dar Zuria (Ethiopia), and Manhiça District (Mozambique) between 20 January 2022 and 24 March 2023. Eligible participants were aged 5–18 years, weighed at least 15 kg, and were infected with *T. trichiura*, hookworm, or *S. stercoralis*; exclusion criteria included benzimidazole treatment within the previous three months and pregnancy. Participants were randomised by block randomisation, stratified by STH species, using sealed envelopes and a randomisation sequence generated in R (version 4.1.2, package blockrand), to receive a single dose of FDC (FDCx1), FDC on three consecutive days (FDCx3), or a single dose of ALB alone. Participants and treatment administrators were not masked to allocation; outcome assessors (laboratory technicians) were masked. The full trial design, eligibility criteria, randomisation procedures, and clinical efficacy and safety outcomes have been reported previously ^12,58^.

For the present microbiome sub-study, we included participants recruited in Ethiopia and Mozambique for whom paired pre-treatment and post-treatment (day 21 ± 7) stool samples were available; we did not include participants recruited in Kenya, as informed consent in that country did not allow samples to be sent out of the country.

### Stool sample collection and STH detection

Participants were provided with stool-collection kits and instructed to collect a fresh stool sample. Stool samples were collected at two time points: at screening, within 7 days before treatment, and at day 21 (± 7 days, range days 14–28) after treatment. Parasitological examinations were conducted according to the ALIVE trial protocol^58^. Briefly, duplicate Kato–Katz thick smears were prepared from each stool sample to detect and quantify *T. trichiura*, *A. lumbricoides* and hookworm eggs^59^; and a modified Baermann method was performed in parallel to detect *S. stercoralis* larvae^60^. All stool samples were processed within 24 h of collection, and Kato–Katz smears were read within 30 min after sample preparation. Aliquots of baseline and post-treatment samples were also analysed by quantitative qPCR for the simultaneous detection of *T. trichiura* and *Schistosoma* spp as previously described^34,61^. Two millilitre aliquots of the same stool samples used for microscopy and qPCR were stored at -80C and sent to the Wellcome Sanger Institute for DNA extraction and metagenomic sequencing.

### Infection intensity, treatment efficacy and adverse event definitions

Infection intensity was expressed as eggs per gram of stool for infections assessed using Kato–Katz factor conversion from egg count under the microscope. Cure was defined as the absence of the corresponding parasite at the post-treatment assessment among participants infected at baseline. Egg reduction rate (ERR) and individual egg reduction rate (ERRi) were calculated from pre- and post-treatment eggs-per-gram counts as previously described^33,62^. For secondary analyses of *T. trichiura*, cure was also defined using qPCR, with a post-treatment cycle-threshold value of at least 50 considered negative, as described previously^34^.

Adverse events were coded using MedDRA Preferred Terms and System Organ Class, and classified as treatment-related based on investigator-assessed causality (possible, probable, or definite)^63^. Gastrointestinal disorders were analysed as a pre-specified subgroup given their plausible relationship with the intestinal microbiome. Only treatment-related adverse events were considered in the association between the microbiome and treatment safety.

### DNA extraction, library preparation and sequencing

Stool samples underwent DNA extraction using the FastDNA Spin Kit for Soil DNA Extraction (MP Biomedicals, Santa Ana, CA, USA), as previously described^64^. Dual-indexed Illumina sequencing libraries were prepared in an automated pipeline using NEBNext Ultra II DNA Library Prep Kit for Illumina reagents and protocols, pooled into a single 81-83 sample multiplex, and sequenced using 150 bp paired-end reads on a single lane of an Illumina NovaSeq 6000. The sequence data for both experiments are available at the ENA BioProject PRJEB79832.

### Sequencing quality control and host read removal

Raw sequencing reads were quality-filtered using fastp v0.23.4 ^65^. Adapter sequences were detected and removed, bases were required to have a Phred quality score of at least 20, and reads were discarded when more than 30% of their bases failed this threshold. Reads containing more than five ambiguous bases or shorter than 50Vbp after trimming were removed as well. Poly-G tails were trimmed and overlapping paired reads were used for base correction. PCR/optical duplicates were removed using Clumpify from BBTools v39.01^66^, with an optical duplicate distance of 12,000, two deduplication passes and no substitutions permitted. Host-derived reads were removed by mapping quality-filtered, deduplicated reads against the human reference genome (GRCh38/hg38) using Bowtie2 v2.5.1^67^ in *--very-sensitive* mode, and unmapped read pairs were retained for downstream analysis. Read counts were tracked at each step (input, post-QC, post-deduplication, post-host-removal; Supplementary Fig. 1). All data from participants with a read count lower than 20 million pairs of reads were excluded.

### Read-based taxonomic profiling

Taxonomic profiles were generated using Sylph v0.8.1^68^. Paired host-depleted reads were converted into sample sketches and profiled against genome sketches constructed from release 226 of GlobDB^69^. Profiling was performed with unknown-sequence correction and a minimum read-sequence identity of 99.5%. Taxonomic assignments were added using sylph-tax v1.2.0^68^ and the corresponding GlobDB taxonomy.

Due to the poorer representation of African microbiomes in public databases^40^, we compared GlobDB release 226 with the Genome Taxonomy Database (GTDB) release 226^70^ using identical Sylph parameters before selecting the reference database. Database performance was evaluated according to the proportion of reads assigned to reference genomes, the proportion remaining unclassified and the number of genomes detected per sample. Although both references reached classification rates over 80% in the vast majority of the samples (Supplementary Fig 1C), per-sample comparison showed that GlobDB left less information unclassified when compared to GTDB (Supplementary Fig 1D). Species-level relative abundances were aggregated at the genus, family and phylum levels as required. Unless otherwise specified, taxonomic analyses were analysed at the phylum, family, genus, and species levels.

With no abundance or prevalence filtering, Sylph-GlobDB detected a total of 6,499 prokaryotic species assigned to 230 families. Of these, 2,848 species and 161 families were detected in both countries, whereas 1,782 species and 45 families were detected only in Mozambique and 1,869 species and 24 families only in Ethiopia.

### Resistome profiling

Antimicrobial resistance genes (ARGs) were detected from host-depleted reads using the Resistance Gene Identifier (RGI) bwt module v6.0.8 and the Comprehensive Antibiotic Resistance Database^71^ (CARD; card data v4.0.1 and prevalence, resistance and variant data v4.0.2). Paired reads were aligned locally against CARD reference sequences. The number of reads assigned to each ARG was obtained from the RGI gene-mapping output by combining completely mapped reads and reads mapped to flanking regions, where applicable. Within each sample, gene mappings supported by fewer than 25 reads were removed. ARG abundances were normalised as reads per kilobase per million host-depleted reads (RPKM). CARD annotations were also used to assign antibiotic drug classes and resistance mechanisms.

### Microbial metabolic pathway profiling

Microbial metabolic pathways were profiled using HUMAnN v3.8^72^. Host-depleted paired reads were concatenated for each sample and analysed using MetaPhlAn v4.0.6^73^ with the mpa_vOct22_CHOCOPhlAnSGB_202212 marker database, together with the full ChocoPhlAn nucleotide and UniRef90 protein databases. Sample-level pathway tables were combined and normalised to copies per million (CPM) as the recommended normalisation approach in HUMAnN ^72^. Unstratified MetaCyc pathway abundances were used to evaluate overall functional diversity and differential abundance. The UNMAPPED and UNINTEGRATED categories were excluded. Species-stratified pathway profiles were retained to identify the microbial taxa contributing to treatment-associated functional changes. For selected pathways, species-level contributions were summarised as mean CPM before and after treatment.

Pathways were manually grouped according to their MetaCyc annotations into amino-acid metabolism, nucleotide and translation-related metabolism, cofactor, quinone and redox metabolism, bacterial cell-wall metabolism, central carbon metabolism and “other” functions.

### Metagenome-assembled genome reconstruction and quality control

Metagenome-assembled genomes (MAGs) were reconstructed independently from the host-depleted paired reads of each sample using mags_maker v1.1.1^74^, a Nextflow implementation of a customized metaWRAP v1.3.2 workflow^75^. QCed reads were assembled using MEGAHIT, and assembled contigs were independently binned using MetaBAT2^76^, MaxBin2^77^ and CONCOCT^78^. The resulting bins were consolidated through the metaWRAP bin-refinement module and subsequently reassembled. Unless otherwise indicated, default workflow parameters were used.

MAG quality was evaluated using qc_mags v1.0.0^79^. The workflow first assigned taxonomy using GTDB-Tk, with the ANI-screening step disabled, and assessed the unprocessed MAGs using QUAST v5.3.0^80^, CheckM2 v1.0.2^81^ and GUNC v1.0.6^82^. Potentially contaminating contigs were then identified and removed using MDMCleaner v0.8.7^83^, after which seqkit v2.10.0^84^ was used to remove contigs below the applicable minimum-length threshold. QUAST, CheckM2 and GUNC were subsequently rerun on the decontaminated MAGs. Taxonomic classification was performed using GTDB release 226.

All downstream analyses used the post-decontamination MAGs and their corresponding quality metrics. MAGs were classified as high quality when they had at least 90% estimated completeness, no more than 5% estimated contamination and passed the GUNC chimerism assessment. These criteria retained 22,032 high-quality MAGs from the 60,738 MAGs initially reconstructed.

### MAG dereplication and taxonomic classification

High-quality MAGs were dereplicated at 95% average nucleotide identity (ANI) using dRep v3.5.0 with fastANI, multiround primary clustering, greedy secondary clustering and tertiary clustering^85^. This produced 1,372 representative species-level genomes. Taxonomic assignments were obtained from GTDB-Tk classifications. Representative genomes that could not be assigned to a named species were retained as poorly characterized or potentially novel species-level populations.

For higher-resolution analysis of treatment-associated changes in *Enterobacteriaceae*, all high-quality MAGs assigned to this family were dereplicated separately at 99% ANI to define strain-level genomic clusters. Cluster prevalence was calculated as the proportion of samples within each country, treatment arm and time point from which a MAG belonging to the corresponding cluster was recovered.

### Phylogenetic analysis of representative MAGs

Representative bacterial and archaeal MAGs were analysed using GTDB-Tk v2.7.1. Conserved bacterial bac120 and archaeal ar53 marker proteins were identified and aligned using the GTDB-Tk identify and align workflows. Approximate maximum-likelihood phylogenies were inferred using FastTree v2.1.10^86^ under the Whelan–Goldman amino-acid substitution model. The fastest search option was additionally applied to the bacterial alignment. Trees were annotated with GTDB taxonomy and country of recovery using ggtree v3.16.3^87^ and ggtreeExtra v1.18.0^88^.

### ARG identification and taxonomic assignment in MAGs

High-quality MAGs were functionally annotated using the merged_annotator workflow in metagenomic mode^89^. ARG annotations from CARD/RGI confirmed by DeepARG at the MAGS level were used as the primary gene-level assignments. To investigate the genomic distribution of ARGs that decreased after FDCx3 treatment, we linked CARD/RGI annotations to the corresponding high-quality MAGs and their GTDB taxonomy. For analyses requiring additional support, CARD-positive MAGs were restricted to genomes that also contained a DeepARG annotation. The prevalence of selected tetracycline-resistance genes—*tet(W)*, *tet(40)*, *tet(O/32/O)* and *tet(O/M/O)*—within each bacterial family was calculated as the number of high-quality MAGs in that family carrying the gene divided by the total number of high-quality MAGs assigned to the family.

### Microbiome diversity analyses

Alpha diversity was quantified using the Shannon index. Between-sample dissimilarity was calculated using Bray–Curtis distances and visualised using principal coordinate analysis (PCoA).

At baseline, we evaluated associations between Shannon diversity and continuous variables, including age, body mass index, and infection intensity, using Spearman correlation. Binary categorical variables were compared using Wilcoxon rank-sum tests, whereas variables with more than two categories were evaluated using Kruskal–Wallis tests. Associations with Bray–Curtis dissimilarity were evaluated using permutational multivariate analysis of variance (PERMANOVA) with 999 permutations. Baseline variables included country, age, sex, body mass index, infecting STH species, infection intensity and the presence of helminth coinfection. Analyses were initially performed in the complete cohort and subsequently stratified by country to reduce confounding by differences in participant characteristics and infection patterns.

To investigate associations between the pretreatment microbiome and treatment efficacy, baseline diversity was compared between cured and non-cured participants, with cure determined by microscopy as the primary outcome. Secondary analyses considered qPCR-defined *T. trichiura* cure and the ERRi which was analysed as a continuous variable. Because cure rates in the FDCx1 and FDCx3 arms were high, cure analyses were restricted to ALB-treated participants with *T. trichiura* infection in Mozambique and ALB-treated participants with hookworm infection in Ethiopia. An exploratory analysis combined the ALB and FDCx1 arms among hookworm-infected Ethiopian participants. Associations with safety were evaluated by comparing the baseline microbiomes of participants who experienced a treatment-related adverse event (AE) with those who did not. Treatment-related gastrointestinal disorders were additionally analysed as a separate outcome. Safety analyses were conducted within each treatment arm and after combining the FDCx1 and FDCx3 arms in an exploratory FDC-level analysis.

Before-and-after changes were evaluated separately within each country and treatment arm by comparing paired pretreatment and post-treatment samples. Changes in Shannon diversity were assessed using paired Wilcoxon signed-rank tests. Differences in Bray–Curtis dissimilarity were evaluated using PERMANOVA with 999 permutations constrained within participant.

P-values from diversity analyses were adjusted using the Bonferroni method within each microbiome profile and analysis family.

### Differential-abundance analyses

Differential abundance was evaluated for bacterial taxa, ARGs and metabolic pathways using MaAsLin3^31^ as the primary method and ANCOM-BC2^32^ as a sensitivity analysis. MaAsLin3 jointly modelled feature prevalence and abundance among samples in which the feature was detected. Taxonomic relative abundances were analysed using total-sum scaling and log transformation, whereas ARG RPKM and pathway CPM profiles were analysed without further normalisation or transformation. Continuous covariates were standardised by MaAsLin3. For ANCOM-BC2 analyses of taxonomic composition, relative abundances were converted to sequencing-depth-scaled pseudo-counts by multiplying each relative abundance by the number of host-depleted reads in the corresponding sample and rounding to the nearest integer. ANCOM-BC2 was run with structural-zero detection disabled and a pseudo-count sensitivity score of zero. For longitudinal analyses, participant identifiers were included as a random effect.

Before differential-abundance testing, taxa were required to have a relative abundance of at least 0.5% and to occur in at least 5% of samples in the corresponding analysis. ARGs and metabolic pathways were required to occur in at least 5% of samples. Features were considered significant in MaAsLin3 at a false-discovery-rate-adjusted joint *q*V<V0.01 and in ANCOM-BC2 at a Bonferroni-adjusted *P*V<V0.05. Features meeting the respective significance thresholds in both methods were considered robustly associated.

Differential-abundance analyses encompassed three sets of comparisons: (1) baseline associations with country, participant characteristics and STH infection variables; (2) associations between the pretreatment microbiome and treatment efficacy or safety; and (3) longitudinal changes between pre- and post-treatment samples. Treatment-outcome analyses used the same clinical outcomes and participant subsets described for the diversity analyses. Baseline infection intensity was included as a covariate in treatment-efficacy models. Longitudinal analyses were performed separately by country and treatment arm, with time point included as the principal explanatory variable and participant identifier as a random effect.

To investigate whether the treatment-associated reduction in *Enterobacteriaceae* could be explained by parasite clearance, longitudinal differential-abundance testing was repeated among ALB-treated participants who achieved parasitological cure. Associations between changes in *Enterobacteriaceae*, *Escherichia* and *E. coli* abundance and the individual doses of ALB and IVM were evaluated using Spearman correlation.

Because requiring statistical significance in both MaAsLin3 and ANCOM-BC2 was conservative, an additional exploratory analysis considered features identified as significant by MaAsLin3 alone at *q*V<V0.01. Significant taxa, ARGs and metabolic pathways were intersected between Ethiopia and Mozambique within each treatment arm to identify cross-country patterns, and between FDCx1 and FDCx3 within each country to identify signals shared by the two FDC regimens.

### Ethical clearance

The ALIVE trial was conducted in accordance with the Declaration of Helsinki and applicable national regulations. All sites approved the trial through their national ethics committees and regulatory authorities. The regulatory goal for the use of the FDC formulations against soil-transmitted helminthiases is subject to review and evaluation by the European Medicines Agency through the EU-Medicines for all procedures (EU-M4all). The trial’s protocol and the primary outcomes of the ALIVE clinical trial have been published^12,58^. The trial was registered at ClinicalTrials.gov under NCT05124691. Written informed consent was obtained from the parent or legal guardian of each participant, and participants aged 12–17 years provided written assent. The informed consent in Ethiopia and Mozambique, as well as the trial protocol, allowed the performance of this microbiome analysis.

## Data availability

The shotgun microbiome sequencing datasets generated in this study have been deposited in the European Nucleotide Archive under BioProject accession <u>PRJEB79832</u>. The 22,032 high-quality metagenome assembled genomes mapped to their 1,372 representative MAGs have been deposited in Zenodo (DOI:10.5281/zenodo.24111237), where we have also provided the Supplementary Tables 12, 13 and 14, that allows the mapping of the ENA raw sequencing file to the samples ID, the MAG ID to the sample ID and the ENA raw sequencing file, and each MAG to its dereplicated representative, respectively.

De-identified individual participant metadata associated with the sequencing files can be shared upon request following publication via the Infectious Diseases Data Observatory. Access will be provided to investigators who submit scientifically rigorous proposals. Each proposal will undergo evaluation for scientific merit by the investigator sponsor and collaborators. Requests for data should be directed to Jose Muñoz. Investigators seeking access to data will need to execute a data access agreement beforehand. After data released, the metadata can be mapped to the sequencing files and the MAGs through the sample ID.

## Code availability

All code is available in the GitHub repository https://github.com/Gandasegui/ALIVE_microbiome.

## Acknowledgements

We thank the children and their families who participated in this study, and the school communities in Bahir Dar, Ethiopia, and Manhiça District, Mozambique, for their support throughout the trial. We thank the Pathogen Informatics team at the Wellcome Sanger Institute for their support with sequencing data management and processing.

## Author information

### Contributions

J.G. and S.R.D. conceived and designed this work. A.K. and J.M. designed and led the ALIVE clinical trial. W.G., A.M.J., D.D., D.B., V.N., Á.d.J., I.M. and W.E. coordinated and conducted trial recruitment, sample collection and clinical data curation at the study sites. J.D., K.A., N.S., A.B. and T.K.J. managed and perform sample shipments, DNA extraction and sequencing. J.G. and P.F. performed the bioinformatic and statistical analyses, with input from Y.S., T.L. and S.R.D. J.G. wrote the original draft of the manuscript, with input from S.R.D. All authors reviewed, edited and approved the final manuscript.

## Extended Data Figures

**Extended Data Fig. 1.**
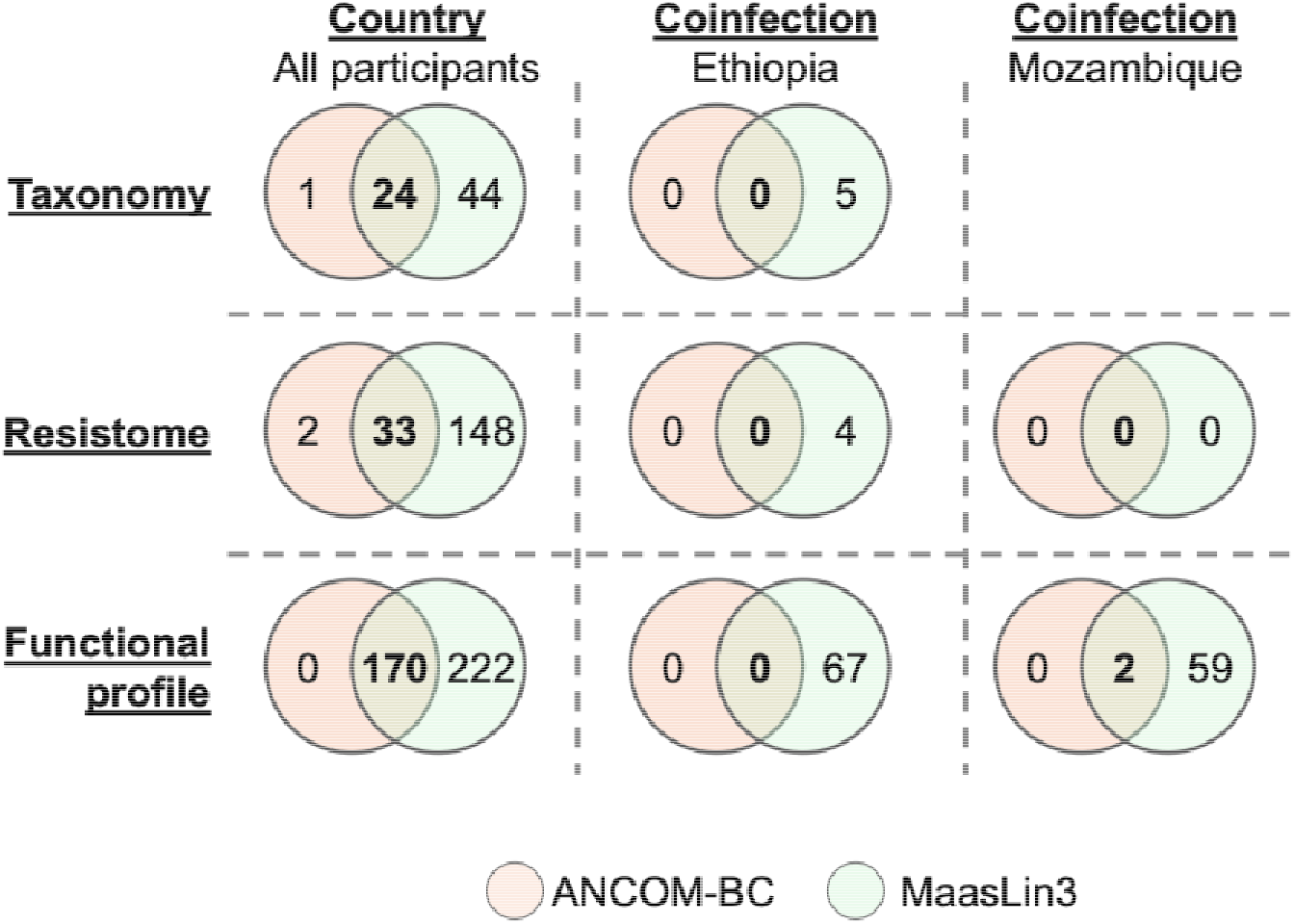
Differential abundance testing of taxonomic, resistome, or functional associations between countries and with coinfection status within countries. Venn diagrams showing the number of features identified as significant by ANCOM-BC2 only, MaAsLin3 only, or both methods (intersection, bold), for taxonomic composition, resistome, and functional profile, comparing country (all participants) and coinfection status within Ethiopia (hookworm-S. stercoralis) and within Mozambique (T. trichiura-A. lumbricoides).

**Extended Data Fig. 2.**
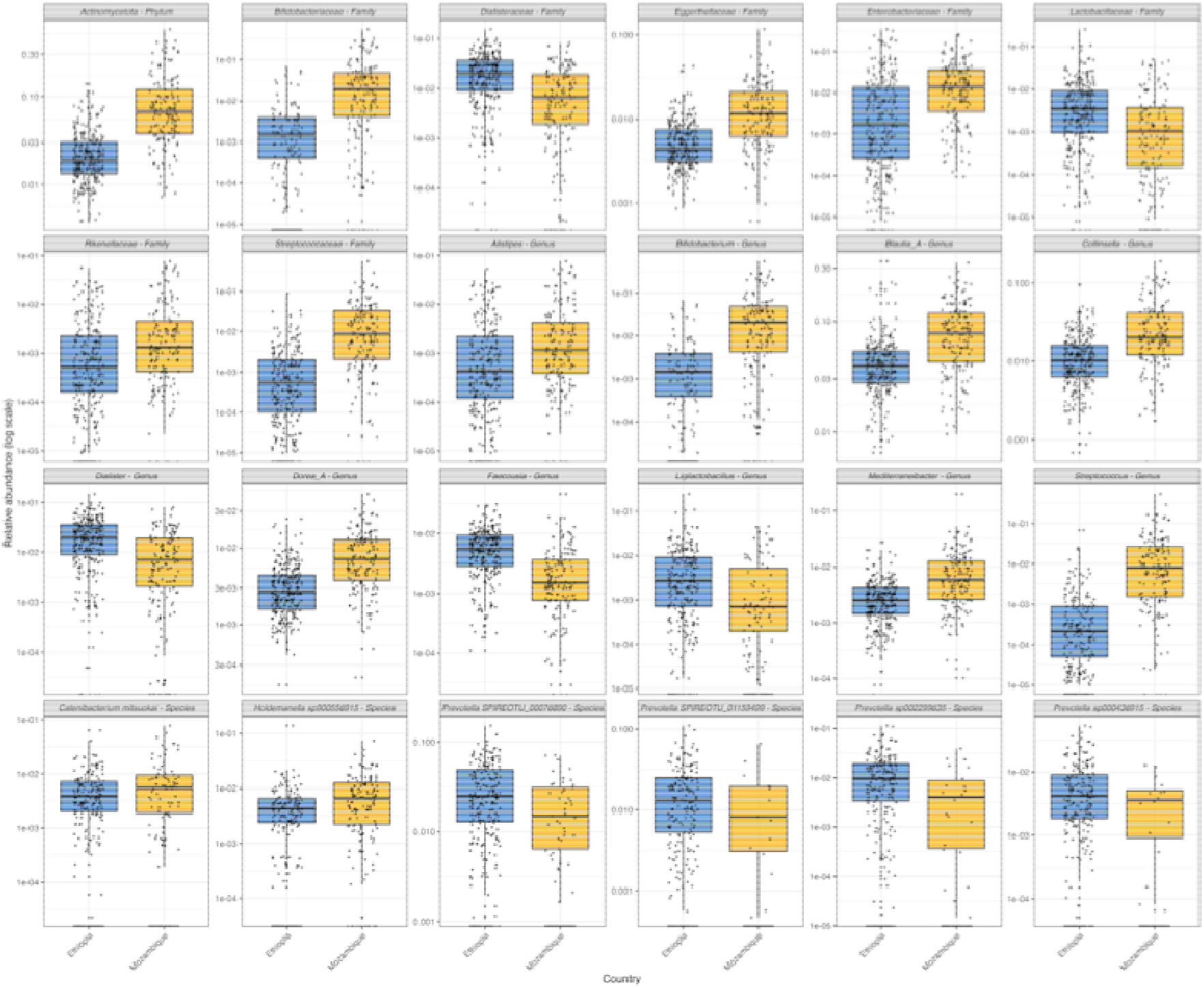
Bacterial taxa differing between the Ethiopian and Mozambican baseline microbiomes. Relative abundances of the 24 taxonomic features identified as significantly different between countries by both MaAsLin3 and ANCOM-BC2. Analyses were performed at the phylum, family, genus and species levels. Points represent individual participants. Relative abundances are displayed on a logarithmic scale.

**Extended Data Fig. 3.**
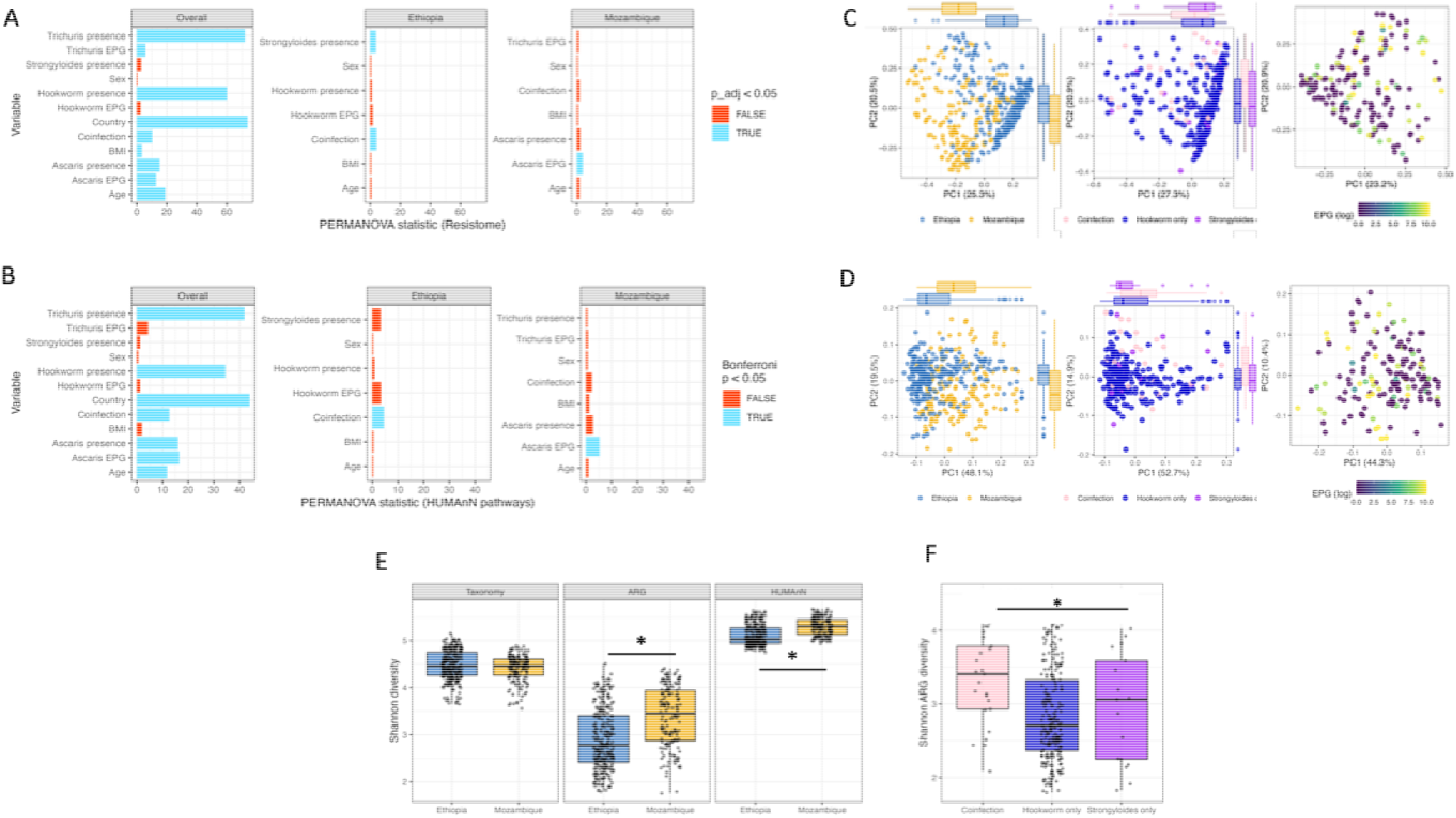
Associations of baseline variables with the resistome and functional profile diversity of the gut microbiome. (A,B) PERMANOVA test statistics for associations between baseline variables and Bray–Curtis dissimilarity of the resistome (A) and HUMAnN metabolic-pathway profile (B), shown for the complete cohort and after stratification by country. (C, D) Principal-coordinate analysis (PCoA) of resistome Bray–Curtis dissimilarity according to country in the complete cohort, helminth infection group in Ethiopia and *Ascaris lumbricoides* infection intensity, expressed as log-transformed eggs per gram of stool (EPG), in Mozambique. (E) Per-country baseline Shannon diversity of the taxonomic, ARG and metabolic-pathway profiles. (F) Baseline ARG Shannon diversity according to helminth infection group in Ethiopia. The asterisk indicates a significant comparison after multiple-testing correction.

**Extended Data Fig. 4.**
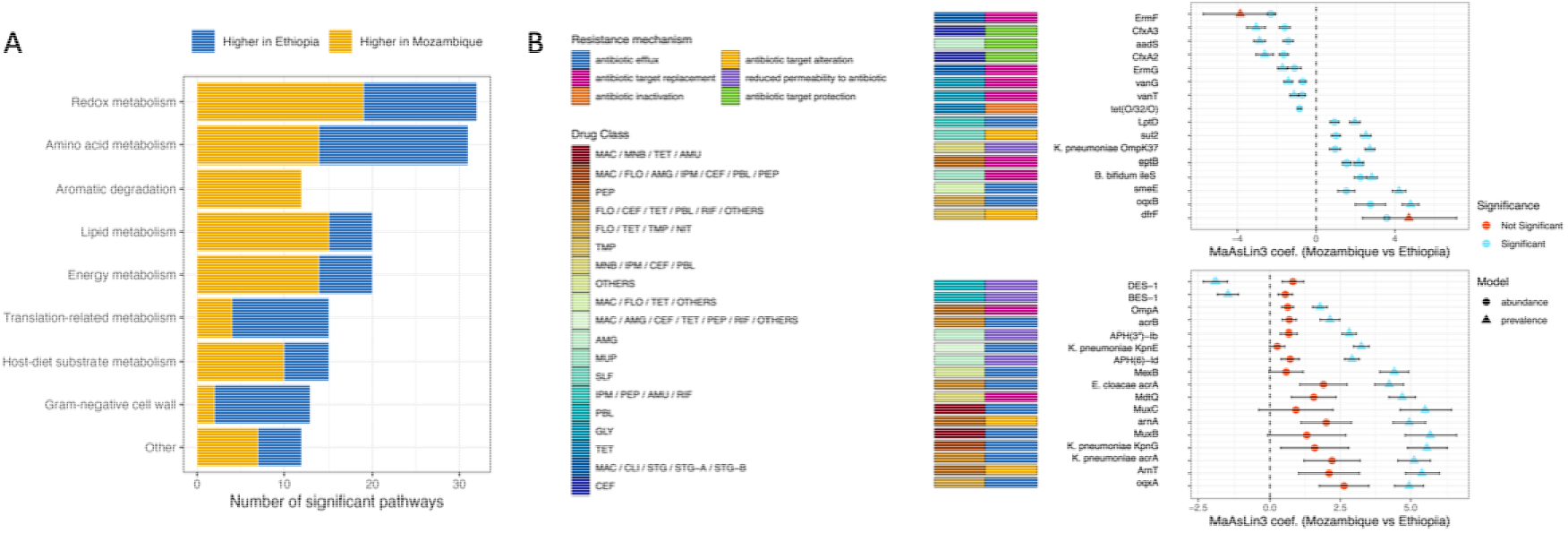
Functional pathways and antibiotic resistance genes (ARGs) differing significantly between Ethiopia and Mozambique at baseline. (A) Number of significant metabolic pathways differing between countries at baseline, grouped by functional category and coloured by the country in which abundance was higher. (B) MaAsLin3 coefficients (Mozambique vs. Ethiopia) for ARGs identified as differentially abundant between countries. Left-hand coloured tiles indicate the resistance mechanism(s) and antibiotic drug class(es) associated with each gene, per the legends shown.

**Extended Data Fig. 5.**
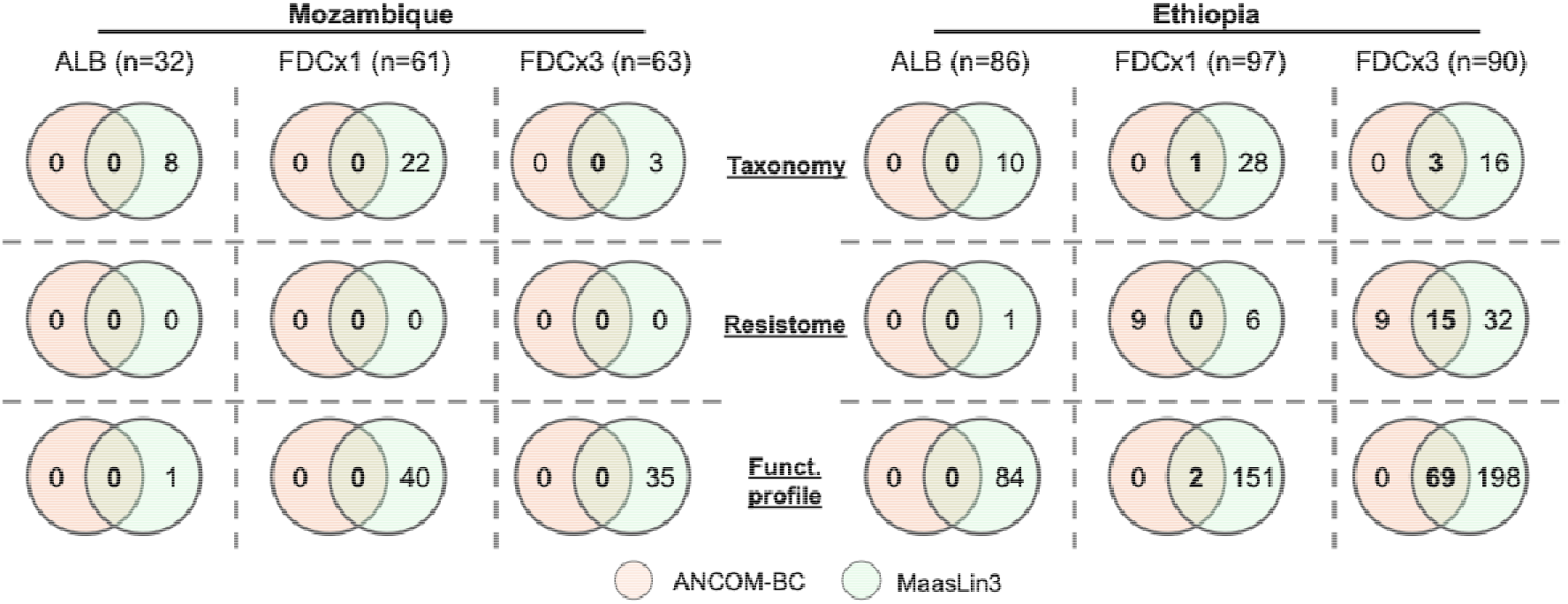
Concordance between differential-abundance methods for treatment-associated microbiome changes. Venn diagrams show the numbers of taxonomic features, antimicrobial resistance genes (ARGs) and metabolic pathways identified as significantly different between pretreatment and post-treatment samples by ANCOM-BC2 and MaAsLin3. Analyses were stratified by country and treatment arm. Intersections represent features identified by both methods. The number of participants included in each paired comparison is indicated above the corresponding column. ALB, albendazole; FDCx1, one dose of the albendazole–ivermectin fixed-dose combination; FDCx3, three consecutive daily doses of the fixed-dose combination.

**Extended Data Fig. 6.**
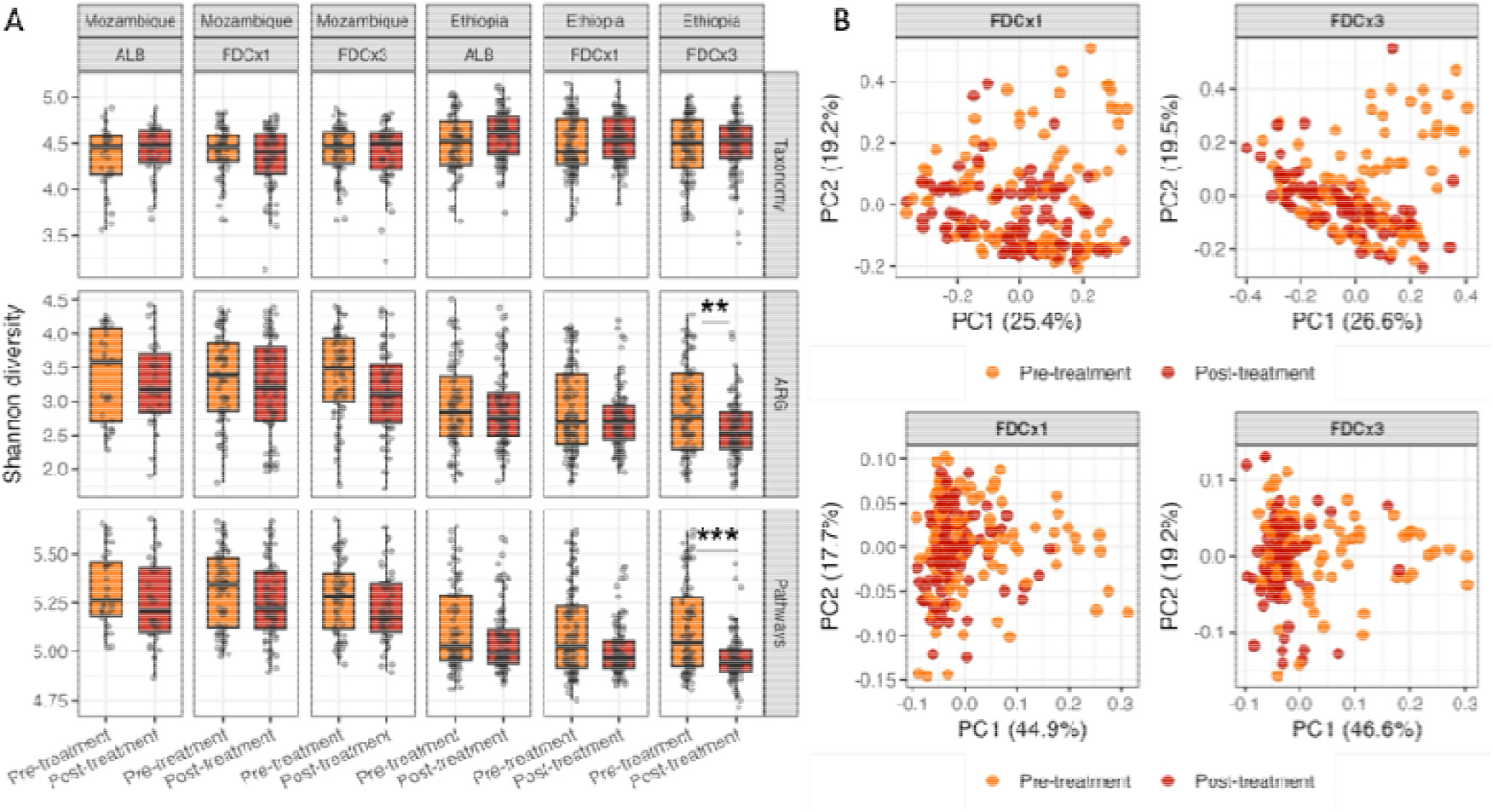
Treatment-associated changes in microbiome diversity. (A) Shannon diversity of the taxonomic, antimicrobial resistance gene (ARG) and metabolic-pathway profiles before and after treatment, stratified by country and treatment arm. Comparisons were performed using paired Wilcoxon signed-rank tests. Asterisks indicate significant paired comparisons after multiple-testing correction; exact adjusted *P* values are provided in Supplementary Table S7. (B) Principal-coordinate analysis of Bray–Curtis dissimilarity for the resistome (top) and metabolic-pathway profile (bottom) before and after FDCx1 or FDCx3 treatment in Ethiopia. Percentages indicate the variation explained by each principal coordinate. ALB, albendazole; FDCx1, one dose of the albendazole–ivermectin fixed-dose combination; FDCx3, three consecutive daily doses of the fixed-dose combination.

**Extended Data Fig. 7.**
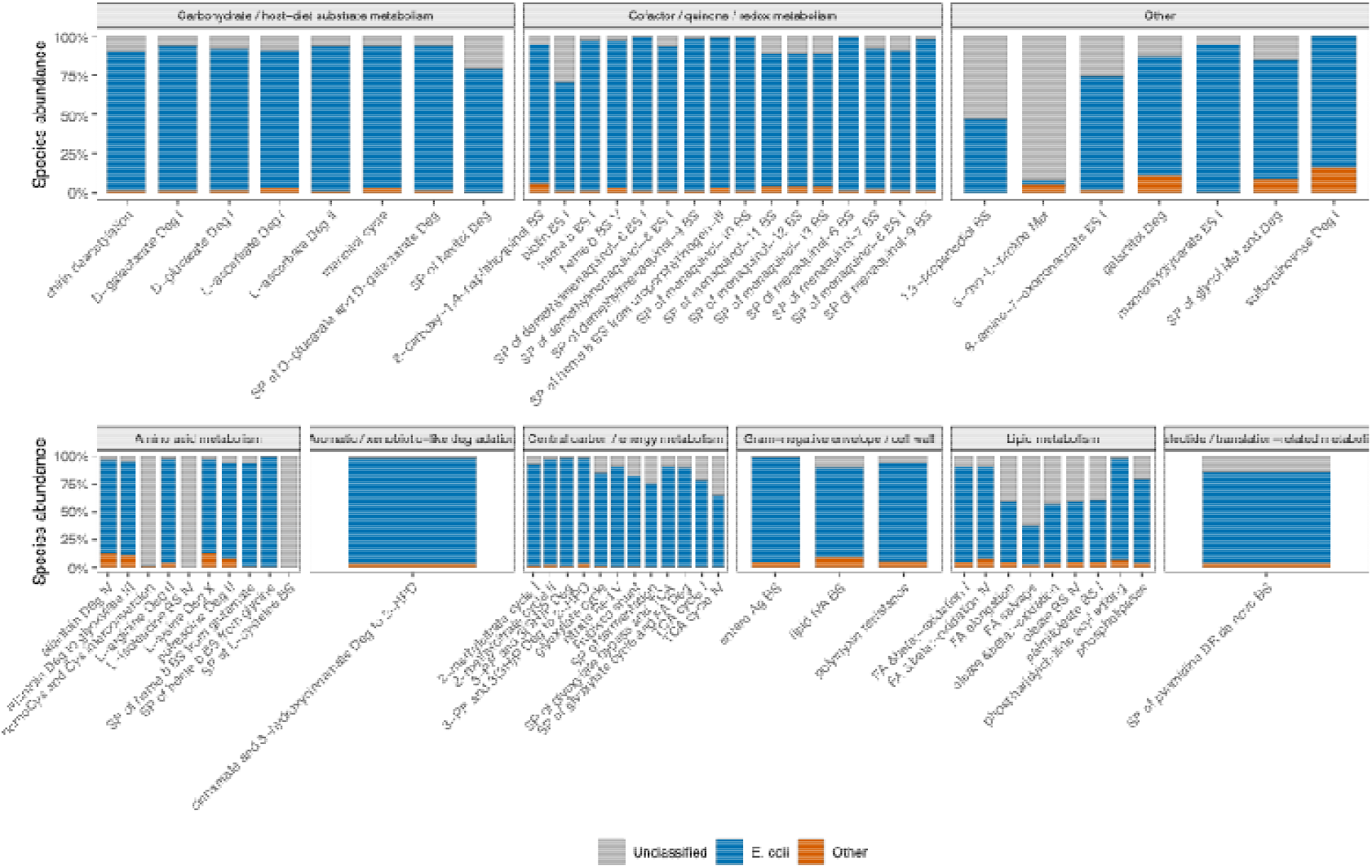
Species contributions to metabolic pathways reduced following FDCx3 treatment in Ethiopia. Species-stratified HUMAnN profiles for the 69 metabolic pathways that decreased significantly after FDCx3 treatment according to both MaAsLin3 and ANCOM-BC2. Pathways are grouped by broad functional theme. Stacked bars show the proportion of pathway abundance attributed to *Escherichia coli*, other classified species or unclassified microbial contributors. BS, biosynthesis; Deg, degradation; FA, fatty acid; SP, superpathway.

**Extended Data Fig. 8.**
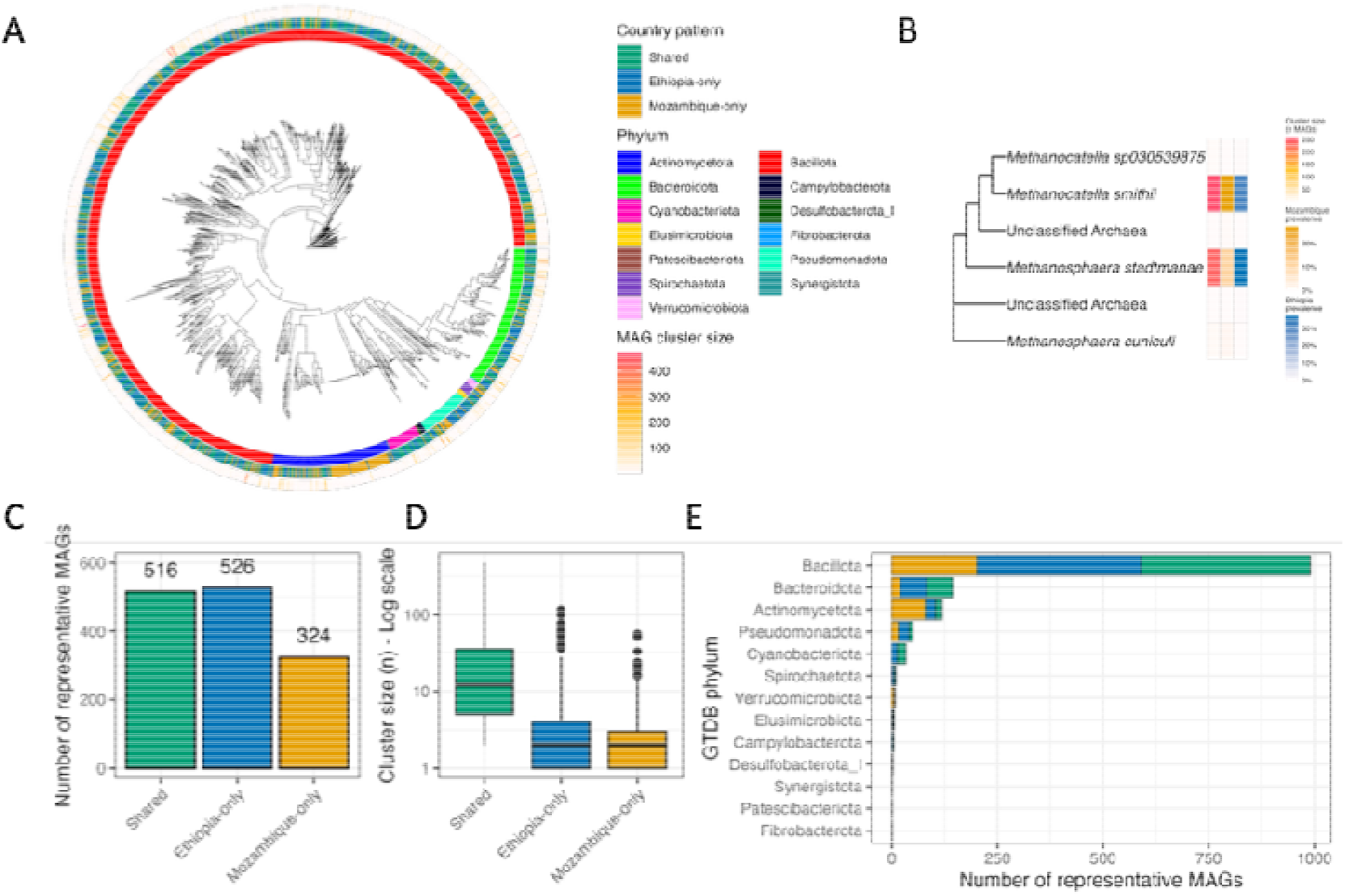
Phylogenetic and geographical distribution of representative metagenome-assembled genomes. (A) Approximate maximum-likelihood phylogeny of the 1,366 representative bacterial metagenome-assembled genomes (MAGs). Concentric annotations indicate GTDB phylum, geographical distribution and species-level cluster size. (B) Phylogeny of the six representative archaeal MAGs. Heat-map annotations show species-level cluster size and MAG recovery prevalence in Ethiopia and Mozambique. (C) Numbers of representative bacterial MAGs recovered in both countries or exclusively in Ethiopia or Mozambique. (D) Distribution of species-level cluster sizes according to geographical distribution, displayed on a logarithmic scale. (E) Number of representative bacterial MAGs within each GTDB phylum, stratified by geographical distribution.

**Extended Data Fig. 9.**
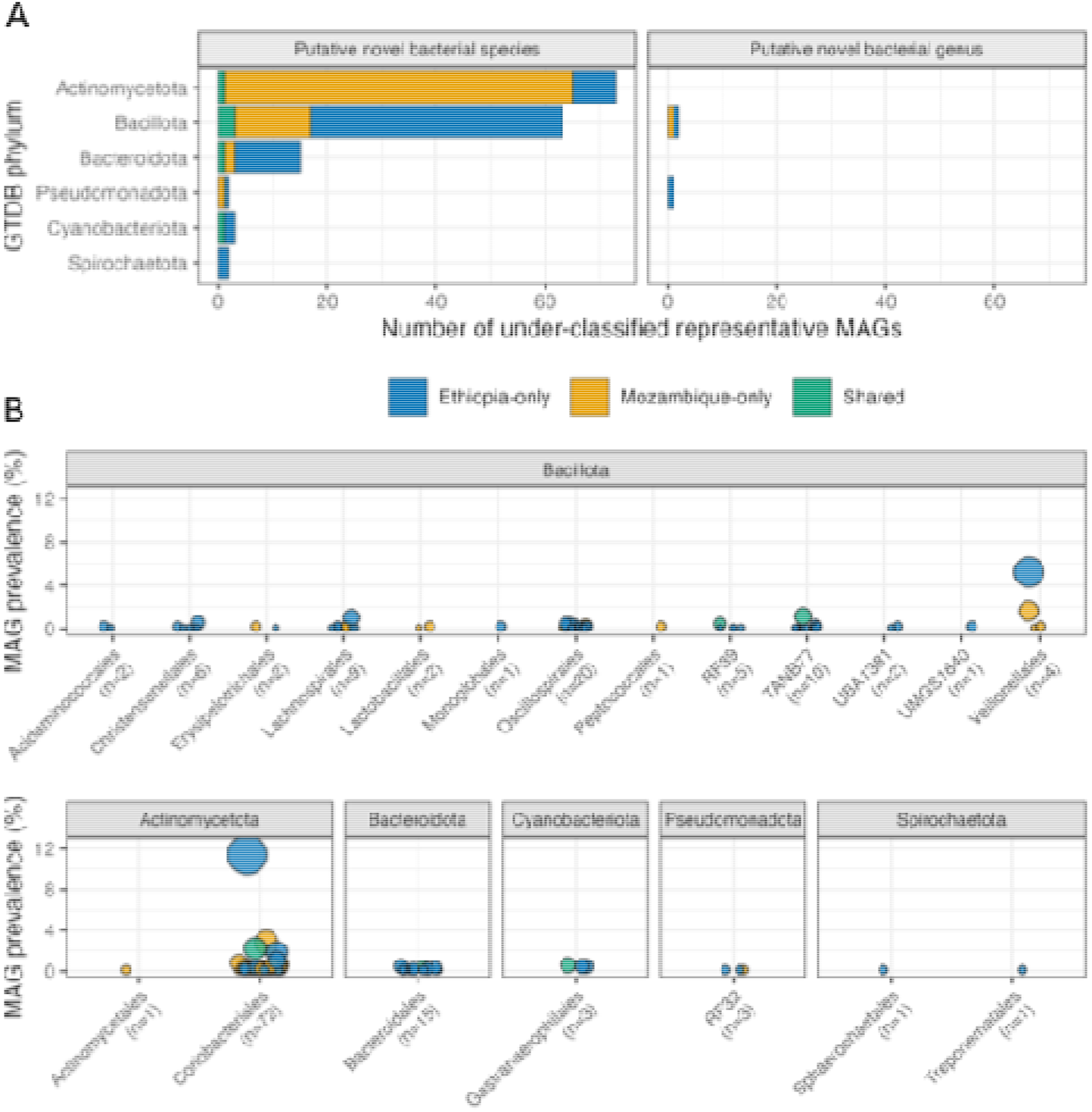
Taxonomic distribution and prevalence of under-classified representative MAGs. (A) Numbers of representative bacterial metagenome-assembled genomes (MAGs) that could not be assigned to a named GTDB species or genus, grouped by GTDB phylum and geographical distribution. (B) Prevalence of under-classified representative MAGs, grouped by GTDB phylum and order. Each point represents one representative genome; point size indicates the number of MAGs within its 95% average-nucleotide-identity cluster. The number of representative genomes within each order is shown in parentheses.

**Extended Data Fig. 10.**
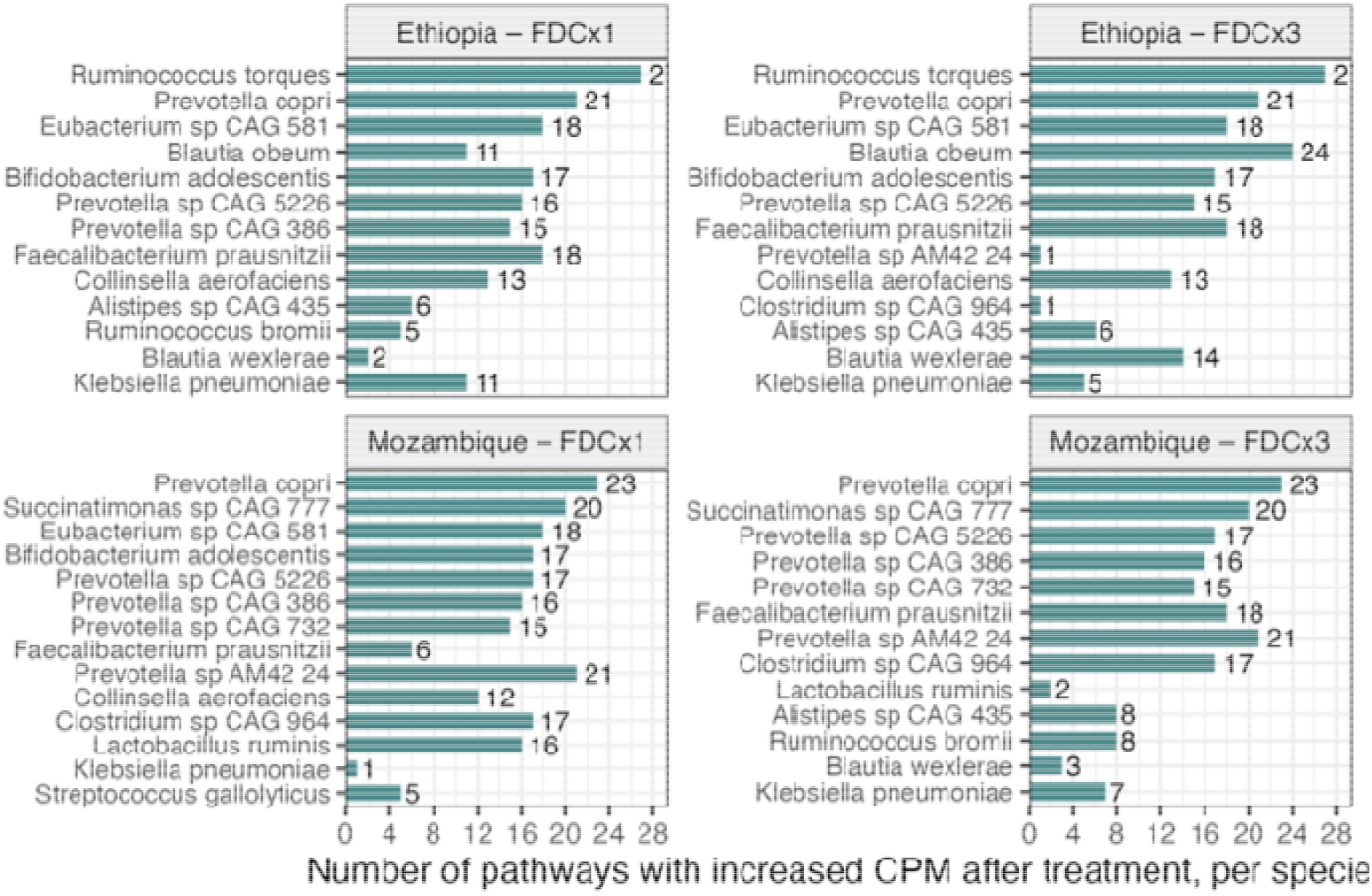
Multiple bacterial species contributed to the increase in pathways shared across FDC comparisons. Numbers of the 28 four-way shared metabolic pathways for which the mean species-stratified contribution increased after treatment, shown separately for each country and FDC regimen. The analysis was restricted to the 20 classified species with the greatest combined pathway abundance among species contributing to at least two pathways. Bar lengths and adjacent numbers indicate the number of pathways with a higher mean species-specific copies-per-million (CPM) abundance after treatment than before treatment. FDCx1, one dose of the albendazole–ivermectin fixed-dose combination; FDCx3, three consecutive daily doses of the fixed-dose combination.

## References

1. Bethony, J. et al. Soil-transmitted helminth infections: ascariasis, trichuriasis, and hookworm. Lancet 367, 1521–1532 (2006).

2. Else, K. J. et al. Whipworm and roundworm infections. Nat. Rev. Dis. Primers 6, 44 (2020).

3. Campbell, S. J., et al. Complexities and perplexities: A critical appraisal of the evidence for soil-transmitted helminth infection-related morbidity. PLoS Negl. Trop. Dis. 10, e0004566 (2016).

4. Yap, P., Utzinger, J., Hattendorf, J. & Steinmann, P. Influence of nutrition on infection and re-infection with soil-transmitted helminths: a systematic review. Parasit. Vectors 7, 229 (2014).

5. Montresor, A. et al. The global progress of soil-transmitted helminthiases control in 2020 and World Health Organization targets for 2030. PLoS Negl. Trop. Dis. 14, e0008505 (2020).

6. Gandasegui, J. et al. Ivermectin and albendazole coadministration: opportunities for strongyloidiasis control. Lancet Infect. Dis. 22, e341–e347 (2022).

7. Emerson, P. M. et al. Need for a paradigm shift in soil-transmitted helminthiasis control: Targeting the right people, in the right place, and with the right drug(s). PLoS Negl. Trop. Dis. 18, e0012521 (2024).

8. Moser, W., Schindler, C. & Keiser, J. Efficacy of recommended drugs against soil transmitted helminths: systematic review and network meta-analysis. BMJ 358, j4307 (2017).

9. Sprecher, V. P. et al. Efficacy and safety of ascending doses of orodispersible ivermectin co-administered with albendazole for Trichuris trichiura infections in preschool-aged children in Tanzania: a single-blind, randomised, controlled, dose-ranging, phase 2 trial. Lancet Infect. Dis. 26, 160–169 (2026).

10. Hürlimann, E. et al. Efficacy and safety of co-administered ivermectin and albendazole in school-aged children and adults infected with Trichuris trichiura in Côte d’Ivoire, Laos, and Pemba Island, Tanzania: a double-blind, parallel-group, phase 3, randomised controlled trial. Lancet Infect. Dis. 22, 123–135 (2022).

11. Matamoros, G. et al. Efficacy and safety of albendazole and high-dose ivermectin coadministration in school-aged children infected with Trichuris trichiura in Honduras: A randomized controlled trial. Clin. Infect. Dis. 73, 1203–1210 (2021).

12. Krolewiecki, A. et al. Albendazole-ivermectin co-formulation for the treatment of Trichuris trichiura and other soil-transmitted helminths: a randomised phase 2/3 trial. Lancet Infect. Dis. 25, 548–559 (2025).

13. Schnoz, A. et al. Efficacy and safety of moxidectin-albendazole combination therapy for Trichuris trichiura infections in school-aged children: a double-blind, randomised, controlled, superiority trial. Lancet Infect. Dis. 25, 1325–1335 (2025).

14. Bolte, L. A., Björk, J. R., Gacesa, R. & Weersma, R. K. Pharmacomicrobiomics: The role of the gut microbiome in immunomodulation and cancer therapy. Gastroenterology 169, 813–827 (2025).

15. Zimmermann, M., Zimmermann-Kogadeeva, M., Wegmann, R. & Goodman, A. L. Separating host and microbiome contributions to drug pharmacokinetics and toxicity. Science 363, eaat9931 (2019).

16. Rosa, B. A., Supali, T., Gankpala, L. & Others. Differential human gut microbiome assemblages during soil-transmitted helminth infections in Indonesia and Liberia. Microbiome 6, 33 (2018).

17. Rubel, M. A., Abbas, A., Taylor, L. J. & Others. Lifestyle and the presence of helminths is associated with gut microbiome composition in Cameroonians. Genome Biology 21, 122 (2020).

18. Gordon, C. A. et al. Helminths, polyparasitism, and the gut microbiome in the Philippines. International Journal for Parasitology 50, 217–225 (2020).

19. Taye, B., Mekonnen, Z., Belanger, K. D. & Davenport, E. R. Gut-microbiome profiles among soil-transmitted helminths (STHs) infected Ethiopian children enrolled in the school-based mass deworming program. PLOS Neglected Tropical Diseases 18, e0012485 (2024).

20. Schneeberger, P. H. H., Dommann, J., Rahman, N. & Others. Profound taxonomic and functional gut microbiota alterations associated with trichuriasis: cross-country and country-specific patterns. npj Biofilms and Microbiomes 12, 45 (2026).

21. Kupritz, J., Angelova, A., Nutman, T. B. & Gazzinelli-Guimaraes, P. H. Helminth-induced human gastrointestinal dysbiosis: a systematic review and meta-analysis reveals insights into altered taxon diversity and microbial gradient collapse. mBio 12, e02890–21 (2021).

22. Ngwese, M. M. et al. Infection with gut parasites correlates with gut microbiome diversity across human populations in Africa. Gut Microbes 17, 2587966 (2025).

23. Loke P’ng, P., Lee, S. C. & Oyesola, O. O. Effects of helminths on the human immune response and the microbiome. Mucosal Immunology 15, 1224–1233 (2022).

24. Loke P’ng, P. & Harris, N. L. Networking between helminths, microbes, and mammals. Cell Host & Microbe 31, 464–471 (2023).

25. Gandasegui, J. et al. Baseline gut microbiota diversity and composition and albendazole efficacy in hookworm-infected individuals. Parasites & Vectors 17, 387 (2024).

26. Schneeberger, P. H. H. et al. Different gut microbial communities correlate with efficacy of albendazole-ivermectin against soil-transmitted helminthiases. Nat. Commun. 13, 1063 (2022).

27. Appiah-Twum, F. et al. The effect of single dose albendazole (400 mg) treatment on the human gut microbiome of hookworm-infected Ghanaian individuals. Scientific Reports 13, 11302 (2023).

28. Easton, A. V. et al. The impact of anthelmintic treatment on human gut microbiota based on cross-sectional and pre- and postdeworming comparisons in western Kenya. mBio 10, (2019).

29. Dommann, J. et al. Combined high-quality metagenomics reveals off-target effects of albendazole, ivermectin-albendazole and moxidectin-albendazole on the human gut bacteria. NPJ Biofilms Microbiomes 12, (2026).

30. Dommann, J. et al. Exposure of gut bacterial isolates to the anthelminthic drugs, ivermectin and moxidectin, leads to antibiotic-like phenotypes of growth inhibition and adaptation. Commun. Biol. 7, 1566 (2024).

31. Nickols, W. A. et al. MaAsLin 3: refining and extending generalized multivariable linear models for meta-omic association discovery. Nat. Methods 23, 554–564 (2026).

32. Lin, H. & Peddada, S. D. Multigroup analysis of compositions of microbiomes with covariate adjustments and repeated measures. Nat. Methods 21, 83–91 (2024).

33. Fleitas, P. E. et al. Individual-level analysis of the efficacy of albendazole and albendazole-ivermectin fixed-dose coformulation (FDC) against Trichuris trichiura and hookworms. Acta Trop. 269, 107742 (2025).

34. Fleitas, P. E. et al. Assessing the diagnostic value of qPCR for Trichuris trichiura: sub-analysis of a multi-country clinical trial to determine the efficacy of albendazole compared to an albendazole-ivermectin fixed dose combination. Front. Parasitol. 4, 1679294 (2025).

35. Cantacessi, C. et al. Impact of experimental hookworm infection on the human gut microbiota. J. Infect. Dis. 210, 1431–1434 (2014).

36. Tran, N. T. D. et al. Chronic Strongyloides stercoralis infection increases presence of the Ruminococcus torques group in the gut and alters the microbial proteome. Sci. Rep. 13, 4216 (2023).

37. He, Y. et al. Regional variation limits applications of healthy gut microbiome reference ranges and disease models. Nat. Med. 24, 1532–1535 (2018).

38. Lee, K. A. & Others. Cross-cohort gut microbiome associations with immune checkpoint inhibitor response in advanced melanoma. Nature Medicine 28, 535–544 (2022).

39. Abdill, R. J., Adamowicz, E. M. & Blekhman, R. Public human microbiome data are dominated by highly developed countries. PLoS Biol. 20, e3001536 (2022).

40. Maghini, D. G. et al. Expanding the human gut microbiome atlas of Africa. Nature 638, 718–728 (2025).

41. Brewster, R. et al. Surveying gut microbiome research in Africans: Toward improved diversity and representation. Trends Microbiol. 27, 824–835 (2019).

42. Shyam Prasad, G., Girisham, S. & Reddy, S. M. Microbial transformation of albendazole. Indian J. Exp. Biol. 48, 415–420 (2010).

43. Tee, M. Z. et al. Gut microbiome of helminth-infected indigenous Malaysians is context dependent. Microbiome 10, 214 (2022).

44. Yin, Q., da Silva, A. C., Zorrilla, F. & Others. Ecological dynamics of Enterobacteriaceae in the human gut microbiome across global populations. Nature Microbiology 10, 541–553 (2025).

45. Doan, T., Yan, D., Arzika, A. M. & Others. Mass azithromycin distribution and antibiotic resistance in the gut and nasopharynx: a cluster-randomized trial. Nature Medicine 32, 859–868 (2026).

46. Ramanan, D. et al. Helminth infection promotes colonization resistance via type 2 immunity. Science 352, 608–612 (2016).

47. Zaiss, M. M. & Harris, N. L. Interactions between the intestinal microbiome and helminth parasites. Parasite Immunology 38, 5–11 (2016).

48. Sargun, A., Gerner, R. R., Raffatellu, M. & Nolan, E. M. Harnessing iron acquisition machinery to target Enterobacteriaceae. J. Infect. Dis. 223, S307–S313 (2021).

49. Seyyedabadi, B., Babataheri, S., Naseri, M., Laher, I. & Soraya, H. Ivermectin and non-parasitic disorders: An update. Curr. Opin. Pharmacol. 85, 102574 (2025).

50. Hague, W. E., Gebski, V. J. & Keech, A. C. Recruitment to randomised studies. Med. J. Aust. 178, 579–581 (2003).

51. Welsche, S., et al. Egg excretion patterns of soil-transmitted helminth infections in humans following albendazole-ivermectin and albendazole treatment. PLoS Negl. Trop. Dis. 18, e0012073 (2024).

52. Palleja, A. & Others. Recovery of gut microbiota of healthy adults following antibiotic exposure. Nature Microbiology 3, 1255–1265 (2018).

53. Anthony, W. E. & Others. Acute and persistent effects of commonly used antibiotics on the gut microbiome and resistome in healthy adults. Cell Reports 39, 110649 (2022).

54. Baldanzi, G. & Others. Antibiotic use and gut microbiome composition links from individual-level prescription data of 14,979 individuals. Nature Medicine 32, 1351–1361 (2026).

55. Gandasegui, J. et al. Gut microbiota composition in travellers is associated with faecal lipocalin-2, a mediator of gut inflammation. Front. Cell. Infect. Microbiol. 14, 1387126 (2024).

56. Elmassry, M. M. et al. A meta-analysis of the gut microbiome in inflammatory bowel disease patients identifies disease-associated small molecules. Cell Host Microbe 33, 218–234.e12 (2025).

57. Almeida, A. et al. A new genomic blueprint of the human gut microbiota. Nature 568, 499–504 (2019).

58. Krolewiecki, A. et al. An adaptive phase II/III safety and efficacy randomized controlled trial of single day or three-day fixed-dose albendazole-ivermectin co-formulation versus albendazole for the treatment of Trichuris trichiura and other STH infections. ALIVE trial protocol. Gates Open Res. 6, 62 (2022).

59. Katz, N., Chaves, A. & Pellegrino, J. A simple device for quantitative stool thick-smear technique in Schistosomiasis mansoni. Rev. Inst. Med. Trop. Sao Paulo 14, 397–400 (1972).

60. Gelaye, W. et al. Performance evaluation of Baermann techniques: The quest for developing a microscopy reference standard for the diagnosis of Strongyloides stercoralis. PLoS Negl. Trop. Dis. 15, e0009076 (2021).

61. Kaisar, M. M. M. et al. Improved diagnosis of Trichuris trichiura by using a bead-beating procedure on ethanol preserved stool samples prior to DNA isolation and the performance of multiplex real-time PCR for intestinal parasites. Parasitology 144, 965–974 (2017).

62. Levecke, B., et al. A comparison of the sensitivity and fecal egg counts of the McMaster egg counting and Kato-Katz thick smear methods for soil-transmitted helminths. PLoS Negl. Trop. Dis. 5, e1201 (2011).

63. Ioannidis, J. P. A. et al. Better reporting of harms in randomized trials: an extension of the CONSORT statement. Ann. Intern. Med. 141, 781–788 (2004).

64. Shao, Y. et al. Stunted microbiota and opportunistic pathogen colonization in caesarean-section birth. Nature 574, 117–121 (2019).

65. Chen, S., Zhou, Y., Chen, Y. & Gu, J. fastp: an ultra-fast all-in-one FASTQ preprocessor. Bioinformatics 34, i884–i890 (2018).

66. BBMap. SourceForge https://sourceforge.net/projects/bbmap/ (2022).

67. Langmead, B. & Salzberg, S. L. Fast gapped-read alignment with Bowtie 2. Nat. Methods 9, 357–359 (2012).

68. Shaw, J. & Yu, Y. W. Rapid species-level metagenome profiling and containment estimation with sylph. Nat. Biotechnol. 43, 1348–1359 (2025).

69. Speth, D. R., et al. GlobDB: a comprehensive species-dereplicated microbial genome resource. Bioinform. Adv. 5, vbaf280 (2025).

70. Parks, D. H. et al. GTDB: an ongoing census of bacterial and archaeal diversity through a phylogenetically consistent, rank normalized and complete genome-based taxonomy. Nucleic Acids Res. 50, D785–D794 (2022).

71. Alcock, B. P. et al. CARD 2023: expanded curation, support for machine learning, and resistome prediction at the Comprehensive Antibiotic Resistance Database. Nucleic Acids Res. 51, D690–D699 (2023).

72. Beghini, F. et al. Integrating taxonomic, functional, and strain-level profiling of diverse microbial communities with bioBakery 3. Elife 10, e65088 (2021).

73. Blanco-Míguez, A. et al. Extending and improving metagenomic taxonomic profiling with uncharacterized species using MetaPhlAn 4. Nat. Biotechnol. 41, 1633–1644 (2023).

74. Mags_maker. (Github).

75. Uritskiy, G. V., DiRuggiero, J. & Taylor, J. MetaWRAP-a flexible pipeline for genome-resolved metagenomic data analysis. Microbiome 6, 158 (2018).

76. Kang, D. D. et al. MetaBAT 2: an adaptive binning algorithm for robust and efficient genome reconstruction from metagenome assemblies. PeerJ 7, e7359 (2019).

77. Wu, Y.-W., Simmons, B. A. & Singer, S. W. MaxBin 2.0: an automated binning algorithm to recover genomes from multiple metagenomic datasets. Bioinformatics 32, 605–607 (2016).

78. Alneberg, J. et al. Binning metagenomic contigs by coverage and composition. Nat. Methods 11, 1144–1146 (2014).

79. Qc_mags: QC Pipeline for Metagenome Assembled Genomes (MAGs). (Github).

80. Gurevich, A., Saveliev, V., Vyahhi, N. & Tesler, G. QUAST: quality assessment tool for genome assemblies. Bioinformatics 29, 1072–1075 (2013).

81. Chklovski, A., Parks, D. H., Woodcroft, B. J. & Tyson, G. W. CheckM2: a rapid, scalable and accurate tool for assessing microbial genome quality using machine learning. Nat. Methods 20, 1203–1212 (2023).

82. Orakov, A. et al. GUNC: detection of chimerism and contamination in prokaryotic genomes. Genome Biol. 22, 178 (2021).

83. Vollmers, J., Wiegand, S., Lenk, F. & Kaster, A.-K. How clear is our current view on microbial dark matter? (Re-)assessing public MAG & SAG datasets with MDMcleaner. Nucleic Acids Res. 50, e76 (2022).

84. Shen, W., Le, S., Li, Y. & Hu, F. SeqKit: A cross-platform and ultrafast toolkit for FASTA/Q file manipulation. PLoS One 11, e0163962 (2016).

85. Olm, M. R., Brown, C. T., Brooks, B. & Banfield, J. F. dRep: a tool for fast and accurate genomic comparisons that enables improved genome recovery from metagenomes through de-replication. ISME J. 11, 2864–2868 (2017).

86. Price, M. N., Dehal, P. S. & Arkin, A. P. FastTree 2--approximately maximum-likelihood trees for large alignments. PLoS One 5, e9490 (2010).

87. Yu, G., Smith, D. K., Zhu, H., Guan, Y. & Lam, T. T.-Y. Ggtree: An r package for visualization and annotation of phylogenetic trees with their covariates and other associated data. Methods Ecol. Evol. 8, 28–36 (2017).

88. Xu, S. et al. GgtreeExtra: Compact visualization of richly annotated phylogenetic data. Mol. Biol. Evol. 38, 4039–4042 (2021).

89. Merged_annotator. (Github).

