## Supplementary Fig for "Genome-resolved metagenomics reveals population-dependent gut microbiome responses to albendazole-ivermectin treatment"

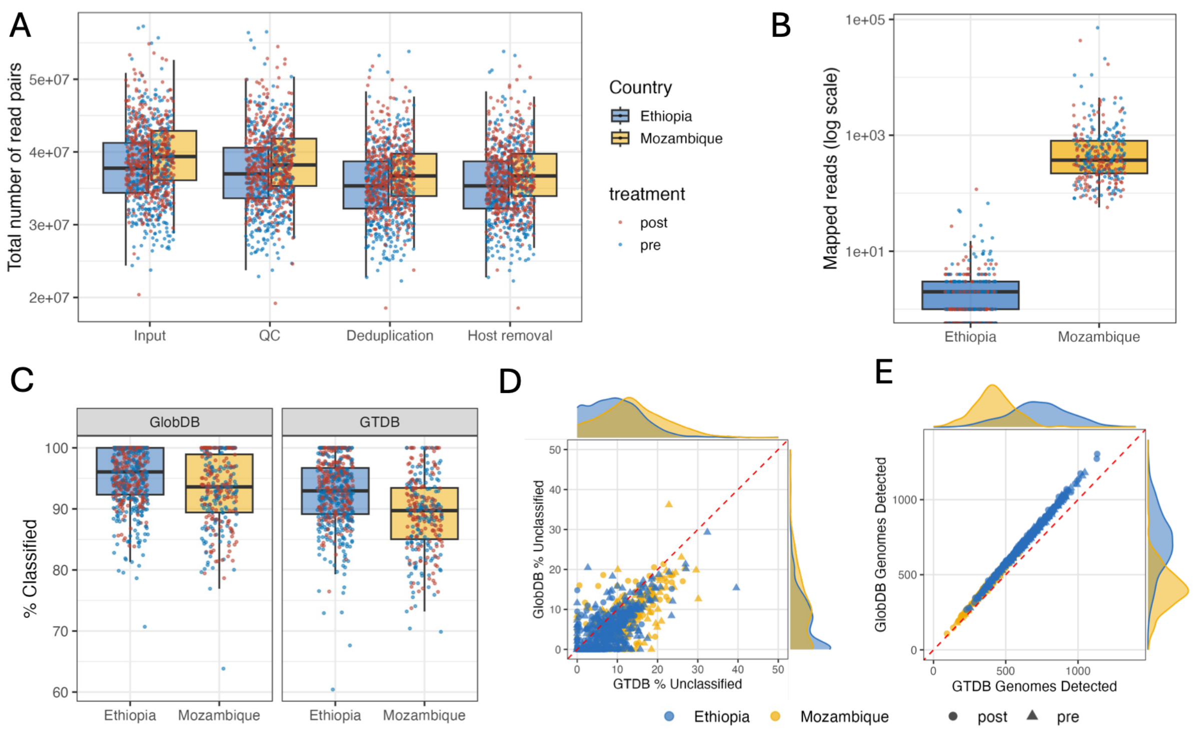


**Supplementary Fig. 1. Sequencing quality control and comparison of GlobDB and GTDB for read-based taxonomic profiling** (A) Total number of read pairs retained at each quality-control step (input, QC, deduplication, host removal), stratified by country (Ethiopia, Mozambique) and coloured by treatment timepoint (pre/post). (B) Number of reads mapped to the human reference genome (log scale) in Ethiopia and Mozambique. (C) Percentage of shotgun metagenomic reads classified using Sylph against GlobDB or GTDB, stratified by country. (B) Percentage of reads remaining unclassified with GlobDB compared with GTDB. (C) Number of reference genomes detected per sample with GlobDB compared with GTDB.

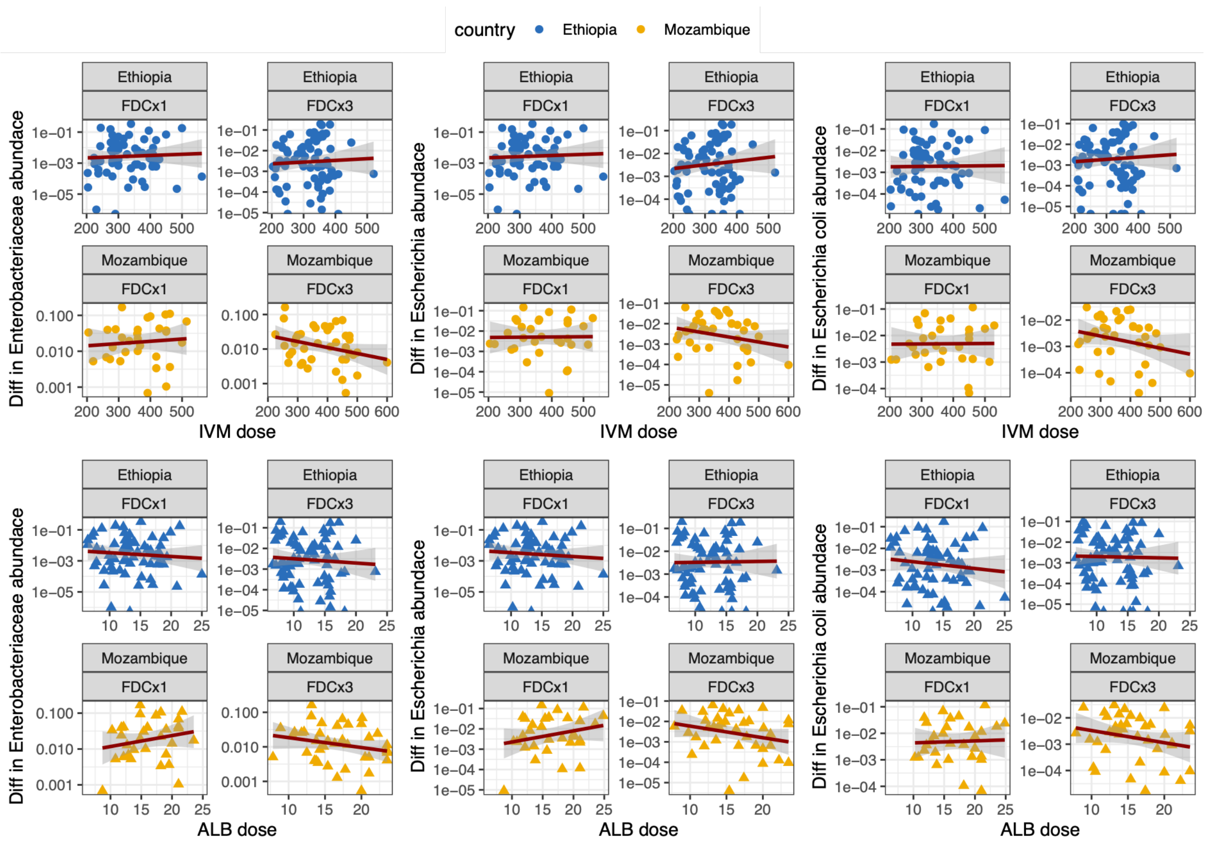


**Supplementary Fig. 2. Associations between individual drug dose and treatment-associated reductions in Enterobacteriaceae.** Associations of individual ivermectin dose (top) and albendazole dose (bottom) with the difference between pretreatment and post-treatment relative abundance of Enterobacteriaceae, Escherichia and Escherichia coli. Analyses are stratified by country and FDC regimen. Ivermectin and albendazole doses are expressed relative to body weight. Points represent individual participants; red lines and shaded areas show linear fits and their 95% confidence intervals. FDCx1, one dose of the albendazole–ivermectin fixed-dose combination; FDCx3, three consecutive daily doses of the fixed-dose combination.

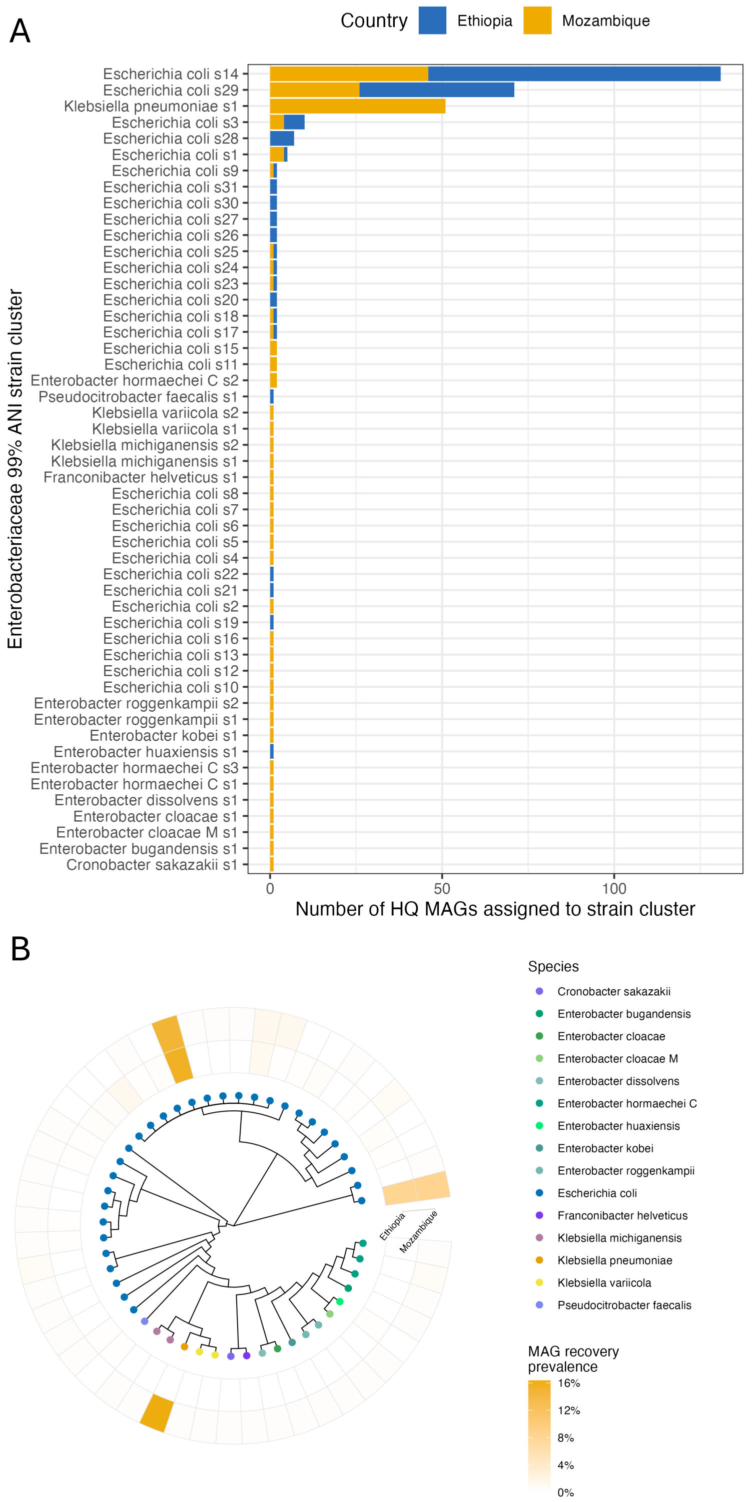


**Supplementary Fig. 3. Genomic diversity and geographical distribution of Enterobacteriaceae strain-level populations.** (A) Number of high-quality Enterobacteriaceae metagenome-assembled genomes (MAGs) assigned to each strain-level cluster. (B) Approximate maximum-likelihood phylogeny of representative Enterobacteriaceae strain-level genomes. HQ, high quality.
